# Air travel-related outbreak of multiple SARS-CoV-2 variants

**DOI:** 10.1101/2021.07.22.21260854

**Authors:** Vijaykrishna Dhanasekaran, Kimberly M Edwards, Ruopeng Xie, Haogao Gu, Dillon C Adam, Lydia DJ Chang, Sammi SY Cheuk, Shreya Gurung, Pavithra Krishnan, Daisy YM Ng, Gigi YZ Liu, Carrie KC Wan, Samuel SM Cheng, Dominic NC Tsang, Benjamin Cowling, Malik Peiris, Leo LM Poon

## Abstract

**Background:** A large cluster of 59 cases were linked to a single flight with 146 passengers from New Delhi to Hong Kong in April 2021. This outbreak coincided with early reports of exponential pandemic growth in New Delhi, which reached a peak of >400,000 newly confirmed cases on 7 May 2021.

**Methods:** Epidemiological information including date of symptom onset, date of positive-sample detection, and travel and contact history for individual cases from this flight were collected. Whole genome sequencing was performed, and sequences were classified based on the dynamic Pango nomenclature system. Maximum-likelihood phylogenetic analysis compared sequences from this flight alongside other cases imported from India to Hong Kong on 26 flights between June 2020 and April 2021, as well as sequences from India or associated with India-related travel from February to April 2021, and 1,217 reference sequences.

**Results:** Sequence analysis identified six lineages of SARS-CoV-2 belonging to two variants of concern (Alpha and Delta) and one variant of public health interest (Kappa) involved in this outbreak. Phylogenetic analysis confirmed at least three independent sub-lineages of Alpha with limited onward transmission, a superspreading event comprising 37 cases of Kappa, and transmission of Delta to only one passenger. Additional analysis of another 26 flights from India to Hong Kong confirmed widespread circulation of all three variants in India since early March 2021.

**Conclusions:** The broad spectrum of disease severity and long incubation period of SARS-CoV-2 pose a challenge for surveillance and control. As illustrated by this particular outbreak, opportunistic infections of SARS-CoV-2 can occur irrespective of variant lineage, and requiring a nucleic acid test within 72 hours of departure may be insufficient to prevent importation or in-flight transmission.

## Introduction

Since March 2020, increasingly stringent travel measures were implemented in Hong Kong to reduce the risk of SARS-CoV-2 importation^1, 2^, including the testing pre-departure, on-arrival, and post-arrival, and quarantine of all passenger arrivals outside the home^3^. Detection of cases on arrival or during quarantine results in contact tracing and additional testing of contacts. In recognition of the emergence of SARS-CoV-2 variants of concern (VOC) with public health significance^4^, on-arrival quarantine was extended from 14 to 21 days on 25 December 2020^5^. Travellers departing from high-risk countries are required to produce a negative nucleic acid test from an accredited laboratory within 72 hours of departure, and passenger flights from countries with widespread VOC circulation are classified as extremely high risk and temporarily banned.

Multiple cases of SARS-CoV-2 were detected upon arrival of a flight from New Delhi to Hong Kong (DEL-HKG). The arrival of this flight in early April 2021 coincided with early reports of exponential growth of SARS-CoV-2 in India^6^ and immediately prior to widespread reports of healthcare infrastructure failure. Because India was at that time classified by the Hong Kong government as high-risk, passengers were required to show documentation of a 21-day quarantine hotel reservation and proof of a negative nucleic acid test taken within 72 hours of departure. After six passengers were denied boarding, the six-hour flight was at 85% capacity. Upon arrival, passengers were again tested by RT-PCR, at which point five positive cases were detected. Cases among passengers continued to be detected throughout the duration of the 21-day quarantine period. For passengers sharing accommodations with positive cases, quarantine periods were extended to 30 days post-arrival. Representing the largest known air travel-related outbreak of SARS-CoV-2^7^, a total of 59 confirmed cases were detected from the 146 passengers onboard, though only 20% (12/59) of cases were reportedly symptomatic. Through whole genome sequencing and phylogenetic analysis, we aimed to infer the dynamics of SARS-CoV-2 transmission associated with this air travel-related related outbreak.

## Methods

Epidemiological information including date of symptom onset, date of positive-sample detection, and travel and contact history for individual cases are provided in Table 1. Of the 59 RT-PCR confirmed SARS-CoV-2 cases, samples from 48 individuals contained adequate viral load for sequencing using previously described methods^8, 9^. Of those, 46 sequences were of sufficient coverage to be classified based on the dynamic Pango nomenclature system (https://pangolin.cog-uk.io/, accessed 10-May-2021)^10^, however those with <70% genome coverage were excluded from subsequent phylogenetic analysis, resulting in 43 complete or nearly complete SARS-CoV-2 genomes. We performed phylogenetic analysis to compare sequences from this outbreak (Table 1) alongside cases imported on 26 additional flights from India to Hong Kong between July 2020 and May 2021 (n=51) (Table 2), as well as GISAID sequences from India or associated with India-related travel from February to April 2021 (n=2,660), and global reference sequences (n=1,217) including Wuhan-Hu-1 (MN908947.3) (accessed 20-May-2021) (Table S1). After removing outliers based on a root-to-tip regression analysis in TempEst v.1.5.3^11^, a maximum likelihood (ML) tree was generated for the final dataset (n=3,971) in IQ-TREE v.2^12, 13^ and dated using the least square dating (LSD2) method^14^.

**Table 1.** SARS-CoV-2 cases detected from a New Delhi to Hong Kong flight in April 2021.

| Case | Travel group | Symptomatic/<br>Asymptomatic | Detection (days<br>post arrival) | Genome<br>coverage* | Pango<br>lineage† |
| --- | --- | --- | --- | --- | --- |
| 1 | Solitary | Asymptomatic | 1 | 99.26 | B.1.617.2 |
| 2 | Solitary | Asymptomatic | 1 | 99.30 | B.1.617.1 |
| 3 | Solitary | Symptomatic | 1 | 99.25 | B.1.617.1 |
| 4 | Solitary | Asymptomatic | 1 | 99.20 | B.1.1.7 |
| 5 | A | Asymptomatic | 1 | 98.15 | B.1.1.7 |
| 6 | A | Asymptomatic | 2 | - | - |
| 7 | A | Symptomatic | 3 | 99.30 | B.1.617.1 |
| 8 | Solitary | Symptomatic | 5 | - | - |
| 9 | B | Asymptomatic | 5 | 99.22 | B.1.1.7 |
| 10 | C | Symptomatic | 6 | 99.30 | B.1.617.1 |
| 11 | B | Symptomatic | 7 | 99.30 | B.1.617.1 |
| 12 | B | Symptomatic | 7 | 99.25 | B.1.617.1 |
| 13 | B | Asymptomatic | 8 | 99.30 | B.1.617.1 |
| 14 | B | Asymptomatic | 8 | 99.30 | B.1.617.1 |
| 15 | B | Asymptomatic | 8 | 99.30 | B.1.617.1 |
| 16 | Solitary | Symptomatic | 8 | 99.25 | B.1.617.1 |
| 17 | Solitary | Symptomatic | 8 | - | - |
| 18 | D | Asymptomatic | 10 | 99.30 | B.1.617.1 |
| 19 | E | Asymptomatic | 11 | 95.08 | B.1.617.1 |
| 20 | D | Asymptomatic | 12 | 99.30 | B.1.617.1 |
| 21 | D | Asymptomatic | 12 | 99.30 | B.1.617.1 |
| 22 | E | Asymptomatic | 13 | 99.30 | B.1.617.1 |
| 23 | F | Asymptomatic | 13 | 95.64 | B.1.617.2 |
| 24 | G | Asymptomatic | 13 | 99.25 | B.1.617.1 |
| 25 | G | Asymptomatic | 13 | 92.17 | B.1.617.1 |
| 26 | H | Asymptomatic | 13 | 99.16 | B.1.617.1 |
| 27 | E | Asymptomatic | 14 | - | - |
| 28 | C | Symptomatic | 14 | 99.30 | B.1.617.1 |
| 29 | D | Asymptomatic | 14 | 95.81 | B.1.617.1 |
| 30 | I | Asymptomatic | 14 | 94.83 | B.1.617.1 |
| 31 | I | Asymptomatic | 14 | 87.34 | B.1.617.1 |
| 32 | I | Asymptomatic | 14 | 99.30 | B.1.617.1 |
| 33 | J | Asymptomatic | 14 | 99.25 | B.1.617.1 |
| 34 | J | Asymptomatic | 14 | 99.30 | B.1.617.1 |
| 35 | Solitary | Asymptomatic | 14 | - | - |
| 36 | K | Asymptomatic | 14 | 96.86 | B.1.617.1 |
| 37 | K | Asymptomatic | 14 | 98.63 | B.1.617.1 |
| 38 | L | Asymptomatic | 14 | 99.30 | B.1.617.1 |
| 39 | L | Asymptomatic | 14 | 72.64 | B.1.617.1 |
| 40 | Solitary | Asymptomatic | 14 | 0.08 | - |
| 41 | H | Asymptomatic | 14 | 96.52 | B.1.617.1 |
| 42 | M | Asymptomatic | 14 | 72.51 | B.1.617.1 |
| 43 | J | Asymptomatic | 14 | 99.30 | B.1.617.1 |
| 44 | Solitary | Asymptomatic | 14 | 92.90 | B.1.617.1 |
| 45 | N | Asymptomatic | 14 | 99.30 | B.1.617.1 |
| 46 | N | Asymptomatic | 14 | 98.61 | B.1.617.1 |
| 47 | O | Symptomatic | 14 | 28.14 | B.1.1.7 |
| 48 | Solitary | Symptomatic | 14 | - | - |
| 49 | O | Asymptomatic | 15 | 45.18 | B.1.1.7 |
| 50 | F | Symptomatic | 15 | 33.95 | B.1.617.1 |
| 51 | M | Asymptomatic | 16 | 99.23 | B.1.617.1 |
| 52 | O | Asymptomatic | 18 | 0.34 | - |
| 53 | P | Asymptomatic | 19 | 99.22 | B.1.617.1 |
| 54 | P | Asymptomatic | 19 | - | - |
| 55 | Solitary | Asymptomatic | 22 | 83.68 | B.1.617.1 |
| 56 | Solitary | Asymptomatic | 23 | - | - |
| 57 | Solitary | Asymptomatic | 24 | - | - |
| 58 | Solitary | Asymptomatic | 26 | - | - |
| 59 | Solitary | Asymptomatic | 30 | - | - |

\*Some samples were unavailable or unable to be sequenced due to insufficient viral load. Genomes with <70% coverage were excluded from phylogenetic analysis.
†Sequences classified according to Pango lineage nomenclature<sup>10</sup>.

**Table 2.** Sequenced SARS-CoV-2 genomes from cases imported to Hong Kong on 26 flights from July 2020 to May 2021.

| Month of arrival | Location of origin | Pango Lineage | Genome coverage (%) | Symptomatic/Asymptomatic |
| --- | --- | --- | --- | --- |
| Jun-20 | Mumbai | B.1 | 96.04 | Asymptomatic |
|  |  | B.1 | 99.3 | Symptomatic |
|  |  | B.1 | 99.3 | Symptomatic |
| Jul-20 | Mumbai via Kuala Lumpur | B.1.210 | 99.3 | Symptomatic |
|  |  | B.1.210 | 99.3 | Symptomatic |
| Jul-20 | New Delhi | B.1.36 | 98.49 | Asymptomatic |
| Jul-20 | India (undetermined location) via Kuala Lumpur | B.1.1 | 99.3 | Symptomatic |
| Jul-20 | Doha via India (undetermined location) | B.1.36.8 | 95.53 | Asymptomatic |
| Aug-20 | New Delhi via Kolkata | B.1.36.36 | 99.3 | Asymptomatic |
| Sep-20 | India (undetermined location) | B.1.36.18 | 99.3 | Asymptomatic |
| Sep-20 | Delhi | B.1.369 | 99.3 | Asymptomatic |
|  |  | B.1.36 | 99.3 | Asymptomatic |
| Sep-20 | Delhi | B.1.1 | 99.3 | Asymptomatic |
| Oct-20 | Delhi | B.1.36.29 | 99.3 | Asymptomatic |
|  |  | B.1.36 | 99.29 | Asymptomatic |
| Oct-20 | Chennai | B.1.1 | 97.18 | Asymptomatic |
|  |  | B.1.1 | 99.3 | Asymptomatic |
| Oct-20 | Chandigarh via Mumbai | B.1.36 | 99.3 | Asymptomatic |
|  |  | B.1.36 | 99.3 | Asymptomatic |
| Nov-20 | Mumbai | B.1.210 | 90.45 | Symptomatic |
| Nov-20 | New Delhi | B.1.36 | 91.73 | Asymptomatic |
| Dec-20 | Delhi | B.1.36 | 98.99 | Asymptomatic |
|  |  | B.1.36 | 97.86 | Symptomatic |
|  |  | B.1.36 | 97.85 | Asymptomatic |
|  |  | B.1.36 | 93.06 | Asymptomatic |
| Dec-20 | Sri Lanka via Mumbai | B.1.1 | 83.21 | Asymptomatic |
| Jan-21 | Mumbai | B.1 | 73.89 | Symptomatic |
|  |  | B.1.1.306 | 99.3 | Symptomatic |
| Feb-21 | Dubai via Bangkok and India (undetermined location) | B.1.562 | 99.02 | Asymptomatic |
| Mar-21 | Kolkata | B.1.1.7 | 99.25 | Asymptomatic |
| Mar-21 | New Delhi | B.1.36.29 | 99.29 | Symptomatic |
| Mar-21 | Kolkata | B.1.1.7 | 99.25 | Symptomatic |
|  |  | B.1.1.7 | 79.79 | Asymptomatic |
|  |  | B.1.1.7 | 91.63 | Asymptomatic |
|  |  | B.1.1.7 | 99.25 | Asymptomatic |
| Mar-21 | Mumbai | B.1.617.2 | 99.28 | Symptomatic |
|  |  | B.1.617.1 | 99.3 | Symptomatic |
| Apr-21 | New Delhi via Dubai | B.1.1.7 | 92.85 | Symptomatic |
| Apr-21 | New Delhi via Kolkata | B.1.617.2 | 99.26 | Asymptomatic |
|  |  | B.1.617.2 | 99.26 | Symptomatic |
|  |  | B.1.617.2 | 99.26 | Asymptomatic |
|  |  | B.1.617.2 | 97.92 | Asymptomatic |
|  |  | B.1.1.7 | 99.2 | Symptomatic |
|  |  | B.1.1.7 | 99.26 | Symptomatic |
|  |  | B.1.617.2 | 99.21 | Symptomatic |
|  |  | B.1.617.2 | 99.26 | Asymptomatic |
| Apr-21 | Doha via India (undetermined location) | B.1.617.2 | 99.26 | Asymptomatic |
| Apr-21 | Mumbai | B.1.617.2 | 88.18 | Asymptomatic |
|  |  | B.1.617.2 | 99.26 | Asymptomatic |
|  |  | B.1.617.1 | 92.9 | Asymptomatic |
|  |  | B.1.617.2 | 92.85 | Asymptomatic |

## Results and Discussion

Whole genome sequencing verified the presence of three distinct variants onboard the DEL-HKG flight, Alpha, Kappa, and Delta^4^, covering three Pango lineages^10^ B.1.1.7, B.1.617.1, and B.1.617.2, respectively. Kappa (B.1.617.1) was detected in the majority of passenger sequences (n=39/46, 84.8%), followed by Alpha (B.1.1.7) in five passengers including a crew member who tested positive on arrival (n=5/46, 10.9%) and Delta (B.1.617.2) in two passengers (n=2/46, 4.3%). In a maximum likelihood phylogeny, 37 of the Kappa sequences clustered together with zero or near-zero branch lengths suggesting a single transmission cluster (inset in Fig. 1A), however one sequence (case 3) was genetically distinct enough to infer pre-flight infection without further onboard transmission. High genome coverage (>98%) of three of the five Alpha variant sequences are consistent with three separate introductions by (i) an infected crew member, (ii) an infected infant, and (iii) an infected but asymptomatic adult in economy seating. Any of these three could represent the index case for onward transmission of the Alpha variant to two individuals from which only partial sequence could be obtained (identified as travel group O in Table 1), testing positive on days 14 and 15 post-arrival. The two Delta variant cases have nearly identical genomes (with >95% coverage), and the two individuals sat in close proximity but tested positive 12 days apart, suggestive of a single onboard transmission event (Table 2 and Fig. 1B). However, there were additional nearby cases which could not be sequenced due to insufficient viral load, and those cases may represent missing links in the inferred transmission chains.

**Figure 1.**
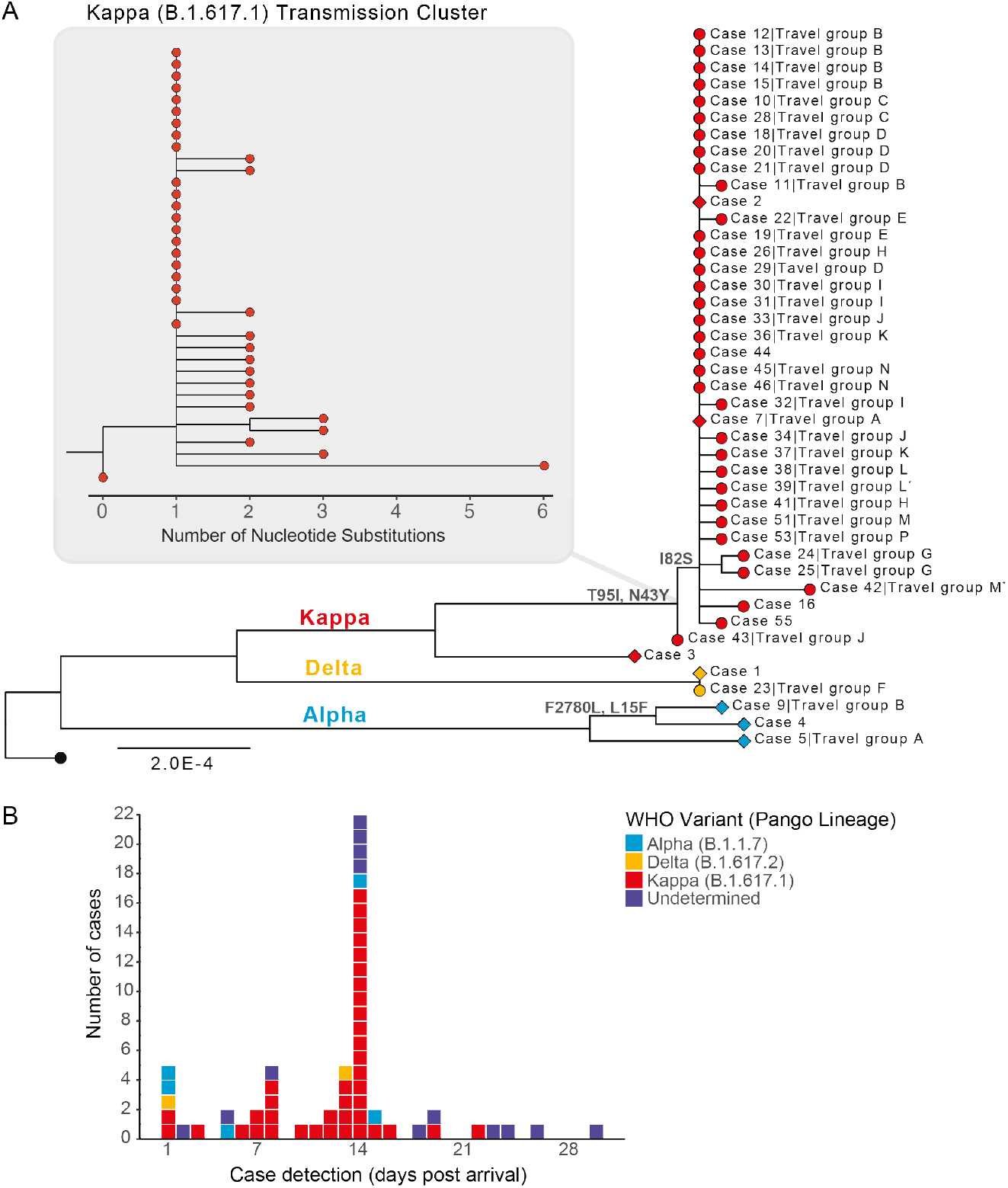
SARS-CoV-2 transmission associated with a flight from New Delhi to Hong Kong in April 2021. Colors represent WHO VOC/VOI and Pango lineage designations10. (A) Maximum likelihood phylogeny of 43 genomes sequenced from 59 cases. Wuhan-Hu-1 (MN908947.3, black tip) was used to root the tree. Diamond tip shapes indicate index cases and circles represent secondary-transmission cases; tips are labelled with case numbers and travel groups. (*) denotes sequences with lower coverage (∼73%). Nodes are labelled with observed amino acid mutations. The Kappa (B.1.617.1) transmission cluster is magnified to show nucleotide substitutions among samples. (B) Case detection colored by SARS-CoV-2 lineage in the 30 days post-arrival. Please contact the corresponding authors to request access to data on seat assignments of passengers testing positive.

Using the temporal window from arrival-to-detection as a proxy for SARS-CoV-2 incubation, we estimate that seven individual cases were likely infected prior to travel (Fig. 1A and Table 1). Forty one cases were detected between days 5 and 14 post arrival, suggesting they were likely infected in transit. Of the cases infected in transit that were sequenced, 94.1% (n=32/34) were related to a single Kappa variant transmission cluster, demonstrating potential for superspreading of SARS-CoV-2 on congested flights and reinforcing the importance of stringent travel-related testing. Another 11 cases were detected after the 14^th^ day post arrival, of which only five could be sequenced. At least two of these Kappa variant cases were likely infected by co-quarantined family members (cases 51 and 53, detected on days 16 and 19, respectively, Fig. 1A and Table 1). Case 59, detected 30 days post-arrival, may be an example of an unusually long incubation period as this person travelled alone and was the only known case from this flight in their quarantine hotel (Table 1). However, this was one of 13 cases not sequenced, and quarantine hotel information was missing for 15 cases in total (n=15/59, 25.4%). Given that instances of SARS-CoV-2 transmission among epidemiologically unlinked cases during quarantine have been documented^15^, we cannot conclusively determine whether transmission occurred in transit or quarantine.

The age distribution of positive cases ranged from four months to 63 years (median age = 33 years), and notably, eight of the positive cases were detected in children under the age of two, who are generally exempt from masking requirements. Interestingly, members of at least two families were infected with independent transmission lineages, and sequencing revealed two instances of children infected with different variants than their family members (Travel groups A and B in Fig. 1A and Table 1). In a family of three individuals (Travel group A), one parent tested positive for Alpha, and the infant tested positive for Kappa (the other parent tested positive, but no sequence was recovered from this patient). As before, using arrival-to-detection as a proxy for incubation, it is likely that Travel group A was infected several days prior to departure. In a second family group (n=6, Travel group B), one child was infected with Alpha, while the other five family members, two children and three adults, were infected with Kappa. The child who tested positive for the Alpha variant at 5 days post arrival was likely infected prior to departure, while the other five family members tested positive on days 7-8 post arrival, suggesting they were most likely infected in-transit.

As a major travel hub, Hong Kong’s practice of testing inbound passengers has proved a valuable source for sentinel SARS-CoV-2 surveillance and containment. Between June 2020 and April 2021, 51 cases of SARS-CoV-2 were detected on 26 other passenger flights from India to Hong Kong (Table 2)^16^. Among those flights, the second largest air-travel related SARS-CoV-2 cluster (n=8 cases) was detected just five days after the DEL-HKG outbreak on another flight arriving from New Delhi (Table 2 and Supplementary Fig. S1). All imported cases detected since mid-March 2021 belonged to Alpha (n=8), Delta (n=13), or Kappa (n=2) lineages and originated from Kolkata, New Delhi, and Mumbai, reflecting the predominant circulation of these variants across major Indian cities during the 2021 resurgence. Similar to the DEL-HKG outbreak, a majority (n=33/51, 64.7%) of imported cases sequenced from the other 26 flights were asymptomatic.

## Conclusions

The broad spectrum of disease severity and potentially long incubation period of SARS-CoV-2 pose a challenge for surveillance and control. Based on the mean incubation period of 5.7 days for SARS-CoV-2^17^ and that infectiousness peaks prior to symptom onset^18^, requiring a nucleic acid test within 72 hours of departure is clearly insufficient to eliminate the risk of SARS-CoV-2 importation or in-flight transmission. Thus, these results highlight the value of quarantine and testing on arrival to prevent virus introduction into the community. A recent review of in-flight transmission of SARS-CoV-2 shows secondary attack rates are highly variable^7^, though, admittedly so are adherence to and enforcement of mask mandates and distancing policies^19^. Furthermore, as illustrated by this particular outbreak, opportunistic infections of SARS-CoV-2 can occur irrespective of variant lineage.

## Data Availability

Hong Kong SARS-CoV-2 genome sequences and associated metadata is deposited at gisaid.org. All anonymized data and sequence accession numbers are available within the manuscript.

## Author Contributions

V.D. and L.L.M.P. designed the study and were responsible for project supervision. K.M.E, R.X., H.G., D.C.A. and D.N.C.T. collected and analyzed the data. L.D.J.C., S.S.Y.C., S.G., P.K., D.Y.M.N., G.Y.Z.L., C.K.C.W., and S.S.M.C performed genome sequencing. V.D., K.M.E and D.C.A. wrote the original draft, and all authors reviewed and edited the manuscript.

## Acknowledgements

We gratefully acknowledge the staff from the originating laboratories responsible for obtaining the specimens and from the submitting laboratories where the genome data were generated and shared via GISAID (Supplementary Table S1). We acknowledge the technical support provided by colleagues from the Centre for PanorOmic Sciences of the University of Hong Kong. We also acknowledge the Centre for Health Protection of the Department of Health for providing epidemiological data for the study.

## Supplementary Data

**Figure S1**. Time-scaled phylogeny of global SARS-CoV-2 genomes related to a flight from New Delhi to Hong Kong (DEL-HKG) in April 2021. Tips are coloured by DEL-HKG flight (n=43, red); global Pango reference sequences (n=1217, purple); cases imported from India to Hong Kong on 26 flights between July 2020 and May 2021 (n=51, green); and GISAID sequences from cases in India or associated with India-related travel (n=2660, blue).

**Figure S2**. Maximum-likelihood phylogeny of the tree in Figure S1 with branch lengths measured in nucleotide substitutions.

**Table S1**. Acknowledgements table.

