## Supplementary figures and images for "Air travel-related outbreak of multiple SARS-CoV-2 variants"

### Figure S1

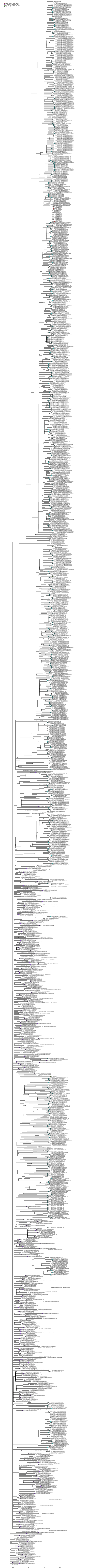

### Figure S2

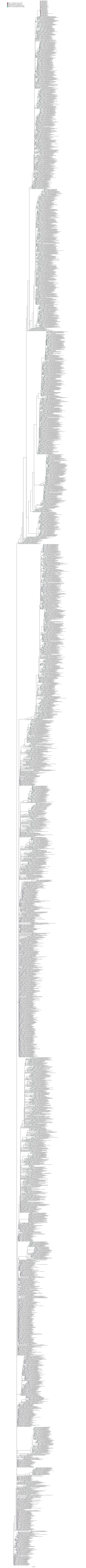
