## Supplementary material for "Air travel-related outbreak of multiple SARS-CoV-2 variants": Table S1

We gratefully acknowledge the following Authors from the Originating laboratories responsible for obtaining the specimens, as well as the Submitting laboratories where the genome data were generated and shared via GISAID, on which this research is based.

All Submitters of data may be contacted directly via www.gisaid.org

Authors are sorted alphabetically.

| Accession ID | Originating Laboratory | Submitting Laboratory | Authors |
|---|---|---|---|
| EPI_ISL_1034266, EPI_ISL_1034270, EPI_ISL_1081924, EPI_ISL_1081930, EPI_ISL_1081945, EPI_ISL_1081946, EPI_ISL_1098834, EPI_ISL_1164353 | National Public Health Laboratory, National Centre for Infectious Diseases | National Public Health Laboratory, National Centre for Infectious Diseases | Tze Minn Mak, Zhenyang Zhou, Lin Cui, Raymond Tzer Pin Lin |
| EPI_ISL_1173251 | National Public Health Laboratory, National Centre for Infectious Diseases | National Public Health Laboratory, National Centre for Infectious Diseases | Tze Minn Mak, Zhenyang Zhou, Royce Ang, Lin Cui, Raymond Tzer Pin Lin |
| EPI_ISL_1229167, EPI_ISL_1252452, EPI_ISL_1252453, EPI_ISL_1295939, EPI_ISL_1312382, EPI_ISL_1312383 | National Public Health Laboratory, National Centre for Infectious Diseases | National Public Health Laboratory, National Centre for Infectious Diseases | Tze Minn Mak, Zhenyang Zhou, Grace Jie Yin Ngan, Royce Ang, Lin Cui, Raymond Tzer Pin Lin |
| EPI_ISL_1357691, EPI_ISL_1357692, EPI_ISL_1357693, EPI_ISL_1357695, EPI_ISL_1357697, EPI_ISL_1357698, EPI_ISL_1357699, EPI_ISL_1357700, EPI_ISL_1357702, EPI_ISL_1357703, EPI_ISL_1357705, EPI_ISL_1357706 | | | |
| see above | INSACOG-WB | National Institute of Biomedical Genomics | Arindam Maitra, Bhaswati Bandyopadhyay, Nidhan Kumar Biswas, Tamal Ghosh, Sreedhar Chinnaswamy, Ajay Chakraborti, Saumitra Das |
| EPI_ISL_1360354, EPI_ISL_1360355 | CSIR-National Environmental Engineering Research Institute | CSIR-Centre for Cellular and Molecular Biology - INSACOG | Ara Sreenivas, Onkar Kulkarni, Lamuk Zaveri, Sofia Banu, Shreekant Verma, Amareshwar Vodapalli , Viswagithe S L, B Himasri, Sharath Chandra Thota, Karthik Bharadwaj Tallapaka, Krishna Khairnar, Rakesh K Mishra,Divya Tej Sowpati |
| EPI_ISL_1360356 | CSIR-National Environmental Engineering Research Institute | CSIR-Centre for Cellular and Molecular Biology - INSACOG | Sofia Banu, Lamuk Zaveri, Ara Sreenivas, Shreekant Verma, Onkar Kulkarni, Amareshwar Vodapalli ,Viswagithe S L, B Himasri, Sharath Chandra Thota, Karthik Bharadwaj Tallapaka, Krishna Khairnar, Rakesh K Mishra,Divya Tej Sowpati |
| EPI_ISL_1360359 | CSIR-National Environmental Engineering Research Institute | CSIR-Centre for Cellular and Molecular Biology - INSACOG | Lamuk Zaveri, Ara Sreenivas, Sofia Banu, Onkar Kulkarni, Shreekant Verma, Amareshwar Vodapalli , Viswagithe S L, B Himasri, Sharath Chandra Thota, Karthik Bharadwaj Tallapaka, Krishna Khairnar, Rakesh K Mishra,Divya Tej Sowpati |
| EPI_ISL_1360360 | CSIR-National Environmental Engineering Research Institute | CSIR-Centre for Cellular and Molecular Biology - INSACOG | Lamuk Zaveri, Ara Sreenivas, Onkar Kulkarni, Sofia Banu, Shreekant Verma,Amareshwar Vodapalli, Viswagithe S L, B Himasri, Sharath Chandra Thota, Karthik Bharadwaj Tallapaka, Krishna Khairnar, Rakesh K Mishra,Divya Tej Sowpati |
| EPI_ISL_1360361, EPI_ISL_1360362 | CSIR-National Environmental Engineering Research Institute | CSIR-Centre for Cellular and Molecular Biology - INSACOG | Ara Sreenivas, Onkar Kulkarni, Lamuk Zaveri, Sofia Banu, Shreekant Verma, Amareshwar Vodapalli , Viswagithe S L, B Himasri, Sharath Chandra Thota, Karthik Bharadwaj Tallapaka, Krishna Khairnar, Rakesh K Mishra,Divya Tej Sowpati |
| EPI_ISL_1360364 | CSIR-National Environmental Engineering Research Institute | CSIR-Centre for Cellular and Molecular Biology - INSACOG | Sofia Banu, Lamuk Zaveri, Ara Sreenivas, Shreekant Verma, Onkar Kulkarni, Amareshwar Vodapalli ,Viswagithe S L, B Himasri, Sharath Chandra Thota, Karthik Bharadwaj Tallapaka, Krishna Khairnar, Rakesh K Mishra,Divya Tej Sowpati |
| EPI_ISL_1360365, EPI_ISL_1360366 | CSIR-National Environmental Engineering Research Institute | CSIR-Centre for Cellular and Molecular Biology - INSACOG | Onkar Kulkarni, Lamuk Zaveri, Ara Sreenivas, Sofia Banu, Shreekant Verma, Amareshwar Vodapalli , Viswagithe S L, B Himasri, Sharath Chandra Thota, Karthik Bharadwaj Tallapaka, Krishna Khairnar, Rakesh K Mishra,Divya Tej Sowpati |
| EPI_ISL_1360367 | CSIR-National Environmental Engineering Research Institute | CSIR-Centre for Cellular and Molecular Biology - INSACOG | Lamuk Zaveri, Ara Sreenivas, Sofia Banu, Onkar Kulkarni, Shreekant Verma, Amareshwar Vodapalli , Viswagithe S L, B Himasri, Sharath Chandra Thota, Karthik Bharadwaj Tallapaka, Krishna Khairnar, Rakesh K Mishra,Divya Tej Sowpati |
| EPI_ISL_1360368 | CSIR-National Environmental Engineering Research Institute | CSIR-Centre for Cellular and Molecular Biology - INSACOG | Lamuk Zaveri, Ara Sreenivas, Onkar Kulkarni, Sofia Banu, Shreekant Verma,Amareshwar Vodapalli, Viswagithe S L, B Himasri, Sharath Chandra Thota, Karthik Bharadwaj Tallapaka, Krishna Khairnar, Rakesh K Mishra,Divya Tej Sowpati |
| EPI_ISL_1360369, EPI_ISL_1360370 | CSIR-National Environmental Engineering Research Institute | CSIR-Centre for Cellular and Molecular Biology - INSACOG | Ara Sreenivas, Onkar Kulkarni, Lamuk Zaveri, Sofia Banu, Shreekant Verma, Amareshwar Vodapalli , Viswagithe S L, B Himasri, Sharath Chandra Thota, Karthik Bharadwaj Tallapaka, Krishna Khairnar, Rakesh K Mishra,Divya Tej Sowpati |
| EPI_ISL_1360371, EPI_ISL_1360372 | CSIR-National Environmental Engineering Research Institute | CSIR-Centre for Cellular and Molecular Biology - INSACOG | Sofia Banu, Lamuk Zaveri, Ara Sreenivas, Shreekant Verma, Onkar Kulkarni, Amareshwar Vodapalli ,Viswagithe S L, B Himasri, Sharath Chandra Thota, Karthik Bharadwaj Tallapaka, Krishna Khairnar, Rakesh K Mishra,Divya Tej Sowpati |
| EPI_ISL_1360373 | CSIR-National Environmental Engineering Research Institute | CSIR-Centre for Cellular and Molecular Biology - INSACOG | Onkar Kulkarni, Lamuk Zaveri, Ara Sreenivas, Sofia Banu, Shreekant Verma, Amareshwar Vodapalli , Viswagithe S L, B Himasri, Sharath Chandra Thota, Karthik Bharadwaj Tallapaka, Krishna Khairnar, Rakesh K Mishra,Divya Tej Sowpati |
| EPI_ISL_1360375 | CSIR-National Environmental Engineering Research Institute | CSIR-Centre for Cellular and Molecular Biology - INSACOG | Lamuk Zaveri, Ara Sreenivas, Sofia Banu, Onkar Kulkarni, Shreekant Verma, Amareshwar Vodapalli , Viswagithe S L, B Himasri, Sharath Chandra Thota, Karthik Bharadwaj Tallapaka, Krishna Khairnar, Rakesh K Mishra,Divya Tej Sowpati |
| EPI_ISL_1360377, EPI_ISL_1360378 | CSIR-National Environmental Engineering Research Institute | CSIR-Centre for Cellular and Molecular Biology - INSACOG | Ara Sreenivas, Onkar Kulkarni, Lamuk Zaveri, Sofia Banu, Shreekant Verma, Amareshwar Vodapalli , Viswagithe S L, B Himasri, Sharath Chandra Thota, Karthik Bharadwaj Tallapaka, Krishna Khairnar, Rakesh K Mishra,Divya Tej Sowpati |
| EPI_ISL_1360381, EPI_ISL_1360382 | CSIR-National Environmental Engineering Research Institute | CSIR-Centre for Cellular and Molecular Biology - INSACOG | Onkar Kulkarni, Lamuk Zaveri, Ara Sreenivas, Sofia Banu, Shreekant Verma, Amareshwar Vodapalli , Viswagithe S L, B Himasri, Sharath Chandra Thota, Karthik Bharadwaj Tallapaka, Krishna Khairnar, Rakesh K Mishra,Divya Tej Sowpati |
| EPI_ISL_1360383 | CSIR-National Environmental Engineering Research Institute | CSIR-Centre for Cellular and Molecular Biology - INSACOG | Lamuk Zaveri, Ara Sreenivas, Sofia Banu, Onkar Kulkarni, Shreekant Verma, Amareshwar Vodapalli , Viswagithe S L, B Himasri, Sharath Chandra Thota, Karthik Bharadwaj Tallapaka, Krishna Khairnar, Rakesh K Mishra,Divya Tej Sowpati |
| EPI_ISL_1360384 | CSIR-National Environmental Engineering Research Institute | CSIR-Centre for Cellular and Molecular Biology - INSACOG | Lamuk Zaveri, Ara Sreenivas, Onkar Kulkarni, Sofia Banu, Shreekant Verma,Amareshwar Vodapalli, Viswagithe S L, B Himasri, Sharath Chandra Thota, Karthik Bharadwaj Tallapaka, Krishna Khairnar, Rakesh K Mishra,Divya Tej Sowpati |
| EPI_ISL_1360385 | CSIR-National Environmental Engineering Research Institute | CSIR-Centre for Cellular and Molecular Biology - INSACOG | Ara Sreenivas, Onkar Kulkarni, Lamuk Zaveri, Sofia Banu, Shreekant Verma, Amareshwar Vodapalli , Viswagithe S L, B Himasri, Sharath Chandra Thota, Karthik Bharadwaj Tallapaka, Krishna Khairnar, Rakesh K Mishra,Divya Tej Sowpati |
| EPI_ISL_1360387, EPI_ISL_1360388 | CSIR-National Environmental Engineering Research Institute | CSIR-Centre for Cellular and Molecular Biology - INSACOG | Sofia Banu, Lamuk Zaveri, Ara Sreenivas, Shreekant Verma, Onkar Kulkarni, Amareshwar Vodapalli ,Viswagithe S L, B Himasri, Sharath Chandra Thota, Karthik Bharadwaj Tallapaka, Krishna Khairnar, Rakesh K Mishra,Divya Tej Sowpati |
| EPI_ISL_1367555, EPI_ISL_1367560 | National Public Health Laboratory, National Centre for Infectious Diseases | National Public Health Laboratory, National Centre for Infectious Diseases | Tze Minn Mak, Zhenyang Zhou, Grace Jie Yin Ngan, Royce Ang, Lin Cui, Raymond Tzer Pin Lin |
| EPI_ISL_1372009 | Patna Medical College, Patna | Institute of Life Sciences - INSACOG | Sunil K. Raghav, Safal Walia, Arup Ghosh, Atimukta Jha, Amol M. Kanampalliwar, Shifu Aggarwal, Rupesh Dash, Rajeeb Swain, Punit Prasad, INSACOG Consortium, Ajay Parida |
| EPI_ISL_1372010 | Immunogenomics lab, Institute of Life Sciences, Bhubaneswar | Institute of Life Sciences - INSACOG | Sunil K. Raghav, Safal Walia, Arup Ghosh, Atimukta Jha, Amol M. Kanampalliwar, Shifu Aggarwal, Rupesh Dash, Rajeeb Swain, Punit Prasad, INSACOG Consortium, Ajay Parida |
| EPI_ISL_1372095 | Nalanda Medical College Hospital, Patna | Institute of Life Sciences - INSACOG | Sunil K. Raghav, Safal Walia, Arup Ghosh, Atimukta Jha, Amol M. Kanampalliwar, Shifu Aggarwal, Rupesh Dash, Rajeeb Swain, Punit Prasad, INSACOG Consortium, Ajay Parida |
| EPI_ISL_1372278 | Mapmygenome | CSIR-Centre for Cellular and Molecular Biology - INSACOG | Ara Sreenivas, Onkar Kulkarni, Lamuk Zaveri, Sofia Banu, Shreekant Verma, Amareshwar Vodapalli , Viswagithe S L, B Himasri, Sharath Chandra Thota, Karthik Bharadwaj Tallapaka, Rakesh K Mishra,Divya Tej Sowpati |
| EPI_ISL_1384818 | National Centre For Cell Science | National Centre For Cell Science | Dhiraj Paul, Sonal Manik Chavan, Mohak P Gujare, Shivang P. Bhanushali, Mitali Inamdar, Manoj Kumar Bhat, Ajay Pillai, Yogesh Shouche |

| | | | |
|---|---|---|---|
| EPI_ISL_1384819, EPI_ISL_1384822, EPI_ISL_1384823, EPI_ISL_1384824, EPI_ISL_1384830, EPI_ISL_1384831, EPI_ISL_1384832, EPI_ISL_1384833, EPI_ISL_1384834, EPI_ISL_1384835, EPI_ISL_1384843, EPI_ISL_1384844, EPI_ISL_1384845, EPI_ISL_1384846, EPI_ISL_1384847, EPI_ISL_1384850, EPI_ISL_1384851, EPI_ISL_1384852 | | | |
| see above | National Centre For Cell Science | National Centre For Cell Science - INSACOG | Dhiraj Paul, Mitali Inamdar, Sonal Manik Chavan, Mohak P Gujare, Shivang P. Bhanushali, Manoj Kumar Bhat, Ajay Pillai, INSACOG Consortium team, Yogesh Shouche. |
| EPI_ISL_1384859 | National Centre For Cell Science | National Institute of Virology, Pune | Dhiraj Paul,Mitali Inamdar, Sonal Manik Chavan, Mohak P Gujare, Shivang P. Bhanushali, Manoj Kumar Bhat, Ajay Pillai, Yogesh Shouche |
| EPI_ISL_1384860, EPI_ISL_1384862, EPI_ISL_1384864, EPI_ISL_1384865, EPI_ISL_1384866, EPI_ISL_1384867, EPI_ISL_1384868, EPI_ISL_1384871, EPI_ISL_1384872, EPI_ISL_1384873, EPI_ISL_1384874, EPI_ISL_1384875, EPI_ISL_1384876, EPI_ISL_1384877, EPI_ISL_1384878, EPI_ISL_1384879, EPI_ISL_1384880, EPI_ISL_1384881, EPI_ISL_1384882, EPI_ISL_1384883, EPI_ISL_1384884 | | | |
| see above | National Centre For Cell Science | National Centre For Cell Science - INSACOG | Dhiraj Paul, Mitali Inamdar, Sonal Manik Chavan, Mohak P Gujare, Shivang P. Bhanushali, Manoj Kumar Bhat, Ajay Pillai, INSACOG Consortium team, Yogesh Shouche. |
| EPI_ISL_1385826, EPI_ISL_1385828 | BBMP Urban PHC | INSACOG-KA, NIMHANS | Chitra Pattabiraman, Pramada Prasad, Anson Kunjumon George, Darshan Sreenivas, Nakka Vijay Kiran Reddy, Anita S Desai, V Ravi |
| EPI_ISL_1415161, EPI_ISL_1415162, EPI_ISL_1415163, EPI_ISL_1415164, EPI_ISL_1415165, EPI_ISL_1415166, EPI_ISL_1415167, EPI_ISL_1415172, EPI_ISL_1415174, EPI_ISL_1415176, EPI_ISL_1415178, EPI_ISL_1415181, EPI_ISL_1415182, EPI_ISL_1415183, EPI_ISL_1415184, EPI_ISL_1415188, EPI_ISL_1415190, EPI_ISL_1415191, EPI_ISL_1415194, EPI_ISL_1415195, EPI_ISL_1415196, EPI_ISL_1415198, EPI_ISL_1415199, EPI_ISL_1415200, EPI_ISL_1415202, EPI_ISL_1415203, EPI_ISL_1415204, EPI_ISL_1415205, EPI_ISL_1415208, EPI_ISL_1415209, EPI_ISL_1415212, EPI_ISL_1415217, EPI_ISL_1415218, EPI_ISL_1415219, EPI_ISL_1415220, EPI_ISL_1415221, EPI_ISL_1415222, EPI_ISL_1415223, EPI_ISL_1415224, EPI_ISL_1415225, EPI_ISL_1415226, EPI_ISL_1415227, EPI_ISL_1415229, EPI_ISL_1415231, EPI_ISL_1415232, EPI_ISL_1415233, EPI_ISL_1415237, EPI_ISL_1415238, EPI_ISL_1415239, EPI_ISL_1415240, EPI_ISL_1415241, EPI_ISL_1415242, EPI_ISL_1415244, EPI_ISL_1415245, EPI_ISL_1415247, EPI_ISL_1415248, EPI_ISL_1415249, EPI_ISL_1415250, EPI_ISL_1415251, EPI_ISL_1415252, EPI_ISL_1415253, EPI_ISL_1415254, EPI_ISL_1415257, EPI_ISL_1415259, EPI_ISL_1415260, EPI_ISL_1415261, EPI_ISL_1415262, EPI_ISL_1415263, EPI_ISL_1415264, EPI_ISL_1415265, EPI_ISL_1415266, EPI_ISL_1415267, EPI_ISL_1415269, EPI_ISL_1415272, EPI_ISL_1415273, EPI_ISL_1415275, EPI_ISL_1415276, EPI_ISL_1415277, EPI_ISL_1415278, EPI_ISL_1415279, EPI_ISL_1415280, EPI_ISL_1415281, EPI_ISL_1415282, EPI_ISL_1415283, EPI_ISL_1415284, EPI_ISL_1415286, EPI_ISL_1415287, EPI_ISL_1415288, EPI_ISL_1415289, EPI_ISL_1415290, EPI_ISL_1415291, EPI_ISL_1415292, EPI_ISL_1415294, EPI_ISL_1415295, EPI_ISL_1415296, EPI_ISL_1415297, EPI_ISL_1415299, EPI_ISL_1415300, EPI_ISL_1415301, EPI_ISL_1415302, EPI_ISL_1415303, EPI_ISL_1415304, EPI_ISL_1415305, EPI_ISL_1415306, EPI_ISL_1415307, EPI_ISL_1415308, EPI_ISL_1415309, EPI_ISL_1415310, EPI_ISL_1415311, EPI_ISL_1415312, EPI_ISL_1415313, EPI_ISL_1415314, EPI_ISL_1415315, EPI_ISL_1415316, EPI_ISL_1415317, EPI_ISL_1415318, EPI_ISL_1415319, EPI_ISL_1415320, EPI_ISL_1415321, EPI_ISL_1415323, EPI_ISL_1415324, EPI_ISL_1415327, EPI_ISL_1415331, EPI_ISL_1415332, EPI_ISL_1415334, EPI_ISL_1415335, EPI_ISL_1415341, EPI_ISL_1415343, EPI_ISL_1415345, EPI_ISL_1415350, EPI_ISL_1415351, EPI_ISL_1415353, EPI_ISL_1415355, EPI_ISL_1415356, EPI_ISL_1415357, EPI_ISL_1415358, EPI_ISL_1415361, EPI_ISL_1415362, EPI_ISL_1415363, EPI_ISL_1415365, EPI_ISL_1415366, EPI_ISL_1415367, EPI_ISL_1415368, EPI_ISL_1415369, EPI_ISL_1415370, EPI_ISL_1415371, EPI_ISL_1415372, EPI_ISL_1415376, EPI_ISL_1415377, EPI_ISL_1415378, EPI_ISL_1415379, EPI_ISL_1415380, EPI_ISL_1415381, EPI_ISL_1415383, EPI_ISL_1415385, EPI_ISL_1415386, EPI_ISL_1415387, EPI_ISL_1415388, EPI_ISL_1415389, EPI_ISL_1415390 | | | |
| see above | National Centre For Cell Science | National Centre For Cell Science - INSACOG | Dhiraj Paul, Mitali Inamdar, Sonal Manik Chavan, Mohak P Gujare, Shivang P. Bhanushali, Manoj Kumar Bhat, Ajay Pillai, INSACOG Consortium team, Yogesh Shouche. |
| EPI_ISL_1419052, EPI_ISL_1419147, EPI_ISL_1419149, EPI_ISL_1419150, EPI_ISL_1419151, EPI_ISL_1419154, EPI_ISL_1419159, EPI_ISL_1419164, EPI_ISL_1419166, EPI_ISL_1419167, EPI_ISL_1419168, EPI_ISL_1419169, EPI_ISL_1419192, EPI_ISL_1419193, EPI_ISL_1419194, EPI_ISL_1419195, EPI_ISL_1419197, EPI_ISL_1419204, EPI_ISL_1419234, EPI_ISL_1419235, EPI_ISL_1419250, EPI_ISL_1419293, EPI_ISL_1419295, EPI_ISL_1419297, EPI_ISL_1419298, EPI_ISL_1419347, EPI_ISL_1419351, EPI_ISL_1419352, EPI_ISL_1419354, EPI_ISL_1419409, EPI_ISL_1419410, EPI_ISL_1419411, EPI_ISL_1419413, EPI_ISL_1419414, EPI_ISL_1419415, EPI_ISL_1419416, EPI_ISL_1419419, EPI_ISL_1419425, EPI_ISL_1419427, EPI_ISL_1419485, EPI_ISL_1419496, EPI_ISL_1419497, EPI_ISL_1419501, EPI_ISL_1419511, EPI_ISL_1419512, EPI_ISL_1419513, EPI_ISL_1419515, EPI_ISL_1419519, EPI_ISL_1419520, EPI_ISL_1419521, EPI_ISL_1419522, EPI_ISL_1419523, EPI_ISL_1419524, EPI_ISL_1419525, EPI_ISL_1419526, EPI_ISL_1419528, EPI_ISL_1419529, EPI_ISL_1419530, EPI_ISL_1419531, EPI_ISL_1419538, EPI_ISL_1419541, EPI_ISL_1419542, EPI_ISL_1419543, EPI_ISL_1419544, EPI_ISL_1419545, EPI_ISL_1419546, EPI_ISL_1419548, EPI_ISL_1419550, EPI_ISL_1419551, EPI_ISL_1419552, EPI_ISL_1419553, EPI_ISL_1419554, EPI_ISL_1419555, EPI_ISL_1419556, EPI_ISL_1419557, EPI_ISL_1419558, EPI_ISL_1419559, EPI_ISL_1419560, EPI_ISL_1419561, EPI_ISL_1419562, EPI_ISL_1419564, EPI_ISL_1419566, EPI_ISL_1419567, EPI_ISL_1419569, EPI_ISL_1419571, EPI_ISL_1419572, EPI_ISL_1419573, EPI_ISL_1419575, EPI_ISL_1419577, EPI_ISL_1419578, EPI_ISL_1419598, EPI_ISL_1419600, EPI_ISL_1419607, EPI_ISL_1419608, EPI_ISL_1419612, EPI_ISL_1419613, EPI_ISL_1419615, EPI_ISL_1419617, EPI_ISL_1419619, EPI_ISL_1419620, EPI_ISL_1419621, EPI_ISL_1419622, EPI_ISL_1419623, EPI_ISL_1419629, EPI_ISL_1419630, EPI_ISL_1419635, EPI_ISL_1419638, EPI_ISL_1419639, EPI_ISL_1419640, EPI_ISL_1419641, EPI_ISL_1419642, EPI_ISL_1419643, EPI_ISL_1419644, EPI_ISL_1419645, EPI_ISL_1419647, EPI_ISL_1419648, EPI_ISL_1419655, EPI_ISL_1419669, EPI_ISL_1419671, EPI_ISL_1419673, EPI_ISL_1419674, EPI_ISL_1419676, EPI_ISL_1419677, EPI_ISL_1419678, EPI_ISL_1419681, EPI_ISL_1419682, EPI_ISL_1419688, EPI_ISL_1419690, EPI_ISL_1419738, EPI_ISL_1419741, EPI_ISL_1419744, EPI_ISL_1419745, EPI_ISL_1419746, EPI_ISL_1419747, EPI_ISL_1419748, EPI_ISL_1419749, EPI_ISL_1419753, EPI_ISL_1419754, EPI_ISL_1419756, EPI_ISL_1419759, EPI_ISL_1419760, EPI_ISL_1419766, EPI_ISL_1419767, EPI_ISL_1419768, EPI_ISL_1419769, EPI_ISL_1419770, EPI_ISL_1419771, EPI_ISL_1419774, EPI_ISL_1419775, EPI_ISL_1419777, EPI_ISL_1419780, EPI_ISL_1419781, EPI_ISL_1419785, EPI_ISL_1419791, EPI_ISL_1419792, EPI_ISL_1419794, EPI_ISL_1419797, EPI_ISL_1419799, EPI_ISL_1419802, EPI_ISL_1419804, EPI_ISL_1419805, EPI_ISL_1419806, EPI_ISL_1419809, EPI_ISL_1419810, EPI_ISL_1419847, EPI_ISL_1419848, EPI_ISL_1419849, EPI_ISL_1419850, EPI_ISL_1419853, EPI_ISL_1419883, EPI_ISL_1419884, EPI_ISL_1419892, EPI_ISL_1419893, EPI_ISL_1419894, EPI_ISL_1419895, EPI_ISL_1419909, EPI_ISL_1419911, EPI_ISL_1419918, EPI_ISL_1419919, EPI_ISL_1419921 | | | |
| see above | INSACOG-WB | National Institute of Biomedical Genomics - INSACOG | Arindam Maitra, Bhaswati Bandyopadhyay, Nidhan Kumar Biswas, Tamal Ghosh, Sreedhar Chinnaswamy, Ajay Chakraborti, Saumitra Das |
| EPI_ISL_1442947, EPI_ISL_1442952, EPI_ISL_1476997, EPI_ISL_1477003, EPI_ISL_1477006, EPI_ISL_1477037, EPI_ISL_1489717, EPI_ISL_1489718, EPI_ISL_1489719, EPI_ISL_1489721, EPI_ISL_1489722, EPI_ISL_1489723, EPI_ISL_1489725, EPI_ISL_1489726 | | | |
| see above | National Public Health Laboratory, National Centre for Infectious Diseases | National Public Health Laboratory, National Centre for Infectious Diseases | Tze Minn Mak, Zhenyang Zhou, Grace Jie Yin Ngan, Royce Ang, Lin Cui, Raymond Tzer Pin Lin |
| EPI_ISL_1511201, EPI_ISL_1511202 | National Centre For Cell Science | National Centre For Cell Science - INSACOG | Dhiraj Paul, Sonal Manik Chavan, Mohak P Gujare, Shivang P. Bhanushali, Mitali Inamdar, Manoj Kumar Bhat, Ajay Pillai, INSACOG Consortium team, Yogesh Shouche |
| EPI_ISL_1519384, EPI_ISL_1519396, EPI_ISL_1519405, EPI_ISL_1519407, EPI_ISL_1519413, EPI_ISL_1519419, EPI_ISL_1524777, EPI_ISL_1524778, EPI_ISL_1524780, EPI_ISL_1524781, EPI_ISL_1524782, EPI_ISL_1524785, EPI_ISL_1524786, EPI_ISL_1524789, EPI_ISL_1524791, EPI_ISL_1524793, EPI_ISL_1524794, EPI_ISL_1524796, EPI_ISL_1524797, EPI_ISL_1524799, EPI_ISL_1524800 | | | |
| see above | National Public Health Laboratory, National Centre for Infectious Diseases | National Public Health Laboratory, National Centre for Infectious Diseases | Tze Minn Mak, Zhenyang Zhou, Grace Jie Yin Ngan, Royce Ang, Lin Cui, Raymond Tzer Pin Lin |
| EPI_ISL_1533738, EPI_ISL_1533742, EPI_ISL_1533743, EPI_ISL_1533744, EPI_ISL_1533745, EPI_ISL_1533746, EPI_ISL_1533747, EPI_ISL_1533748, EPI_ISL_1533750, EPI_ISL_1533752, EPI_ISL_1533753, EPI_ISL_1533754, EPI_ISL_1533755, EPI_ISL_1533756, EPI_ISL_1533758, EPI_ISL_1533762, EPI_ISL_1533763, EPI_ISL_1533771, EPI_ISL_1533778, EPI_ISL_1533780, EPI_ISL_1533783, EPI_ISL_1533784, EPI_ISL_1533785, EPI_ISL_1533790, EPI_ISL_1533791, EPI_ISL_1533792, EPI_ISL_1533793, EPI_ISL_1533794, EPI_ISL_1533795, EPI_ISL_1533796, EPI_ISL_1533798, EPI_ISL_1533799, EPI_ISL_1533800, EPI_ISL_1533801 | | | |
| see above | National Centre For Cell Science | National Centre For Cell Science - INSACOG | Dhiraj Paul, Sonal Manik Chavan, Mohak P Gujare, Shivang P. Bhanushali, Mitali Inamdar, Manoj Kumar Bhat, Ajay Pillai, INSACOG Consortium team, Yogesh Shouche |
| EPI_ISL_1540451, EPI_ISL_1540452, EPI_ISL_1540453, EPI_ISL_1540454, EPI_ISL_1540455, EPI_ISL_1540456, EPI_ISL_1540457 | National Centre For Cell Science | National Centre For Cell Science - INSACOG | Dhiraj Paul, Mohak P Gujare, Shivang P. Bhanushali, Mitali Inamdar, Sonal Manik Chavan, Manoj Kumar Bhat, Ajay Pillai, INSACOG Consortium team, Yogesh Shouche |
| EPI_ISL_1543965, EPI_ISL_1543966, EPI_ISL_1543968, EPI_ISL_1543969, EPI_ISL_1543971, EPI_ISL_1543977, EPI_ISL_1543979, EPI_ISL_1543980, EPI_ISL_1543981 | National Public Health Laboratory, National Centre for Infectious Diseases | National Public Health Laboratory, National Centre for Infectious Diseases | Tze Minn Mak, Zhenyang Zhou, Grace Jie Yin Ngan, Royce Ang, Lin Cui, Raymond Tzer Pin Lin |
| EPI_ISL_1543989, EPI_ISL_1543991, EPI_ISL_1543992, EPI_ISL_1543994, EPI_ISL_1543995, EPI_ISL_1543996, EPI_ISL_1543997, EPI_ISL_1543998, EPI_ISL_1543999, EPI_ISL_1544003, EPI_ISL_1544004, EPI_ISL_1544006, EPI_ISL_1544008, EPI_ISL_1544009, EPI_ISL_1544011, EPI_ISL_1544013, EPI_ISL_1544017, EPI_ISL_1544019, EPI_ISL_1544025, EPI_ISL_1544026, EPI_ISL_1544027, EPI_ISL_1544028, EPI_ISL_1544029, EPI_ISL_1544030, EPI_ISL_1544031, EPI_ISL_1544033, EPI_ISL_1544034, EPI_ISL_1544036, EPI_ISL_1544037, EPI_ISL_1544040, EPI_ISL_1544042, EPI_ISL_1544045, EPI_ISL_1544046, EPI_ISL_1544047, EPI_ISL_1544048, EPI_ISL_1544049, EPI_ISL_1544050, EPI_ISL_1544051, EPI_ISL_1544052, EPI_ISL_1544055, EPI_ISL_1544058, EPI_ISL_1544060, EPI_ISL_1544061, EPI_ISL_1544062, EPI_ISL_1544063, EPI_ISL_1544064, EPI_ISL_1544065, EPI_ISL_1544067, EPI_ISL_1544068, EPI_ISL_1544071 | | | |
| see above | National Centre For Cell Science | National Centre For Cell Science - INSACOG | Dhiraj Paul, Mohak P Gujare, Shivang P. Bhanushali, Mitali Inamdar, Sonal Manik Chavan, Manoj Kumar Bhat, Ajay Pillai, INSACOG Consortium team, Yogesh Shouche |
| EPI_ISL_1544110 | B.J. Medical College and Civil hospital, Ahmedabad | Gujarat Biotechnology Research Centre | Pranay Shah,Zuber Saiyed,Ramesh Pandit,Janvi Raval,Zarna Patel,Nitin Savaliya,Dinesh Kumar,Twinkle Soni,Sonal Sharma ,Kamlesh J Upadhyay,Sanjay Kapadia,Dipa Kinariwala,Umang Mishra,Nitesh Shah,Chaitanya Joshi,Madhvi Joshi |
| EPI_ISL_1544111 | B.J. Medical College and Civil hospital, Ahmedabad | Gujarat Biotechnology Research Centre | Kamlesh J Upadhyay,Ramesh Pandit,Janvi Raval,Zarna Patel,Nitin Savaliya,Dinesh Kumar,Twinkle Soni,Sonal Sharma ,Zuber Saiyed,Pranay Shah,Sanjay Kapadia,Dipa Kinariwala,Umang Mishra,Nitesh Shah,Chaitanya Joshi,Madhvi Joshi |
| EPI_ISL_1544112 | B.J. Medical College and Civil hospital, Ahmedabad | Gujarat Biotechnology Research Centre | Sanjay Kapadia,Janvi Raval,Zarna Patel,Nitin Savaliya,Dinesh Kumar,Twinkle Soni,Sonal Sharma ,Zuber Saiyed,Ramesh Pandit,Pranay Shah,Kamlesh J Upadhyay,Dipa Kinariwala,Umang Mishra,Nitesh Shah,Chaitanya Joshi,Madhvi Joshi |
| EPI_ISL_1544113 | B.J. Medical College and Civil hospital, Ahmedabad | Gujarat Biotechnology Research Centre | Dipa Kinariwala,Zarna Patel,Nitin Savaliya,Dinesh Kumar,Twinkle Soni,Sonal Sharma ,Zuber Saiyed,Ramesh Pandit,Janvi Raval,Pranay Shah,Kamlesh J Upadhyay,Sanjay Kapadia,Umang Mishra,Nitesh Shah,Chaitanya Joshi,Madhvi Joshi |
| EPI_ISL_1544114 | B.J. Medical College and Civil hospital, Ahmedabad | Gujarat Biotechnology Research Centre | Umang Mishra,Nitin Savaliya,Dinesh Kumar,Twinkle Soni,Sonal Sharma ,Zuber Saiyed,Ramesh Pandit,Janvi Raval,Zarna Patel,Pranay Shah,Kamlesh J Upadhyay,Sanjay Kapadia,Dipa Kinariwala,Nitesh Shah,Chaitanya Joshi,Madhvi Joshi |
| EPI_ISL_1544115 | B.J. Medical College and Civil hospital, Ahmedabad | Gujarat Biotechnology Research Centre | Umang Mishra,Dinesh Kumar,Twinkle Soni,Sonal Sharma ,Zuber Saiyed,Ramesh Pandit,Janvi Raval,Zarna Patel,Nitin Savaliya,Pranay Shah,Kamlesh J Upadhyay,Sanjay Kapadia,Dipa Kinariwala,Nitesh Shah,Chaitanya Joshi,Madhvi Joshi |
| EPI_ISL_1544116 | B.J. Medical College and Civil hospital, Ahmedabad | Gujarat Biotechnology Research Centre | Nitesh Shah,Twinkle Soni,Sonal Sharma ,Zuber Saiyed,Ramesh Pandit,Janvi Raval,Zarna Patel,Nitin Savaliya,Dinesh Kumar,Pranay Shah,Kamlesh J |

| | | | |
|---|---|---|---|
| | | | Upadhyay,Sanjay Kapadia,Dipa Kinariwala,Umang Mishra,Chaitanya Joshi,Madhvi Joshi |
| EPI_ISL_1544117 | B.J. Medical College and Civil hospital, Ahmedabad | Gujarat Biotechnology Research Centre | Nitesh Shah,Sonal Sharma ,Zuber Saiyed,Ramesh Pandit,Janvi Raval,Zarna Patel,Nitin Savaliya,Dinesh Kumar,Twinkle Soni,Pranay Shah,Kamlesh J Upadhyay,Sanjay Kapadia,Dipa Kinariwala,Umang Mishra,Chaitanya Joshi,Madhvi Joshi |
| EPI_ISL_1544133 | GMERS, Medical College, Gandhinagar | Gujarat Biotechnology Research Centre | Gaurishankar Shrimali,Sonal Sharma ,Zuber Saiyed,Ramesh Pandit,Janvi Raval,Zarna Patel,Nitin Savaliya,Dinesh Kumar,Twinkle Soni,Umang Mishra,Nitesh Shah,Chaitanya Joshi,Madhvi Joshi |
| EPI_ISL_1544134 | GMERS, Medical College, Gandhinagar | Gujarat Biotechnology Research Centre | Zuber Saiyed,Ramesh Pandit,Janvi Raval,Zarna Patel,Nitin Savaliya,Dinesh Kumar,Twinkle Soni,Sonal Sharma ,Gaurishankar Shrimali,Umang Mishra,Nitesh Shah,Chaitanya Joshi,Madhvi Joshi |
| EPI_ISL_1547802, EPI_ISL_1547803, EPI_ISL_1547804, EPI_ISL_1547805 | CSIR-National Environmental Engineering Research Institute | CSIR-Centre for Cellular and Molecular Biology - INSACOG | Onkar Kulkarni, Lamuk Zaveri, Ara Sreenivas, Sofia Banu, Shreekant Verma, Amareshwar Vodapalli , Viswagithe S L, B Himasri, Sharath Chandra Thota, Karthik Bharadwaj Tallapaka, Krishna Khairnar, Rakesh K Mishra,Divya Tej Sowpati |
| EPI_ISL_1568432 | National Public Health Laboratory, National Centre for Infectious Diseases | National Public Health Laboratory, National Centre for Infectious Diseases | Tze Minn Mak, Zhenyang Zhou, Grace Jie Yin Ngan, Royce Ang, Lin Cui, Raymond Tzer Pin Lin |
| EPI_ISL_1589745, EPI_ISL_1589747, EPI_ISL_1589748, EPI_ISL_1589749, EPI_ISL_1589750, EPI_ISL_1589751, EPI_ISL_1589752, EPI_ISL_1589753, EPI_ISL_1589754, EPI_ISL_1589755, EPI_ISL_1589756, EPI_ISL_1589757, EPI_ISL_1589758, EPI_ISL_1589759, EPI_ISL_1589760, EPI_ISL_1589761, EPI_ISL_1589762, EPI_ISL_1589763, EPI_ISL_1589764, EPI_ISL_1589765, EPI_ISL_1589766, EPI_ISL_1589767, EPI_ISL_1589768, EPI_ISL_1589769, EPI_ISL_1589770, EPI_ISL_1589771, EPI_ISL_1589772, EPI_ISL_1589773, EPI_ISL_1589774, EPI_ISL_1589775, EPI_ISL_1589776, EPI_ISL_1589777, EPI_ISL_1589778, EPI_ISL_1589779, EPI_ISL_1589780, EPI_ISL_1589781, EPI_ISL_1589782, EPI_ISL_1589783, EPI_ISL_1589784, EPI_ISL_1589785, EPI_ISL_1589786, EPI_ISL_1589787, EPI_ISL_1589788, EPI_ISL_1589789, EPI_ISL_1589790, EPI_ISL_1589791, EPI_ISL_1589792, EPI_ISL_1589795, EPI_ISL_1589796, EPI_ISL_1589797, EPI_ISL_1589798, EPI_ISL_1589799, EPI_ISL_1589800, EPI_ISL_1589801, EPI_ISL_1589802, EPI_ISL_1589803, EPI_ISL_1589804, EPI_ISL_1589805, EPI_ISL_1589806, EPI_ISL_1589807, EPI_ISL_1589808, EPI_ISL_1589809, EPI_ISL_1589810, EPI_ISL_1589811, EPI_ISL_1589812, EPI_ISL_1589813, EPI_ISL_1589814, EPI_ISL_1589815, EPI_ISL_1589816, EPI_ISL_1589817, EPI_ISL_1589818, EPI_ISL_1589819, EPI_ISL_1589820, EPI_ISL_1589821, EPI_ISL_1589822, EPI_ISL_1589853, EPI_ISL_1589854, EPI_ISL_1589857, EPI_ISL_1589858, EPI_ISL_1589859, EPI_ISL_1589860, EPI_ISL_1589861, EPI_ISL_1589862, EPI_ISL_1589863, EPI_ISL_1589864, EPI_ISL_1589865, EPI_ISL_1589866, EPI_ISL_1589867, EPI_ISL_1589871, EPI_ISL_1589872, EPI_ISL_1589873, EPI_ISL_1589874, EPI_ISL_1589875, EPI_ISL_1589876, EPI_ISL_1589877, EPI_ISL_1589878, EPI_ISL_1589879, EPI_ISL_1589880, EPI_ISL_1589881, EPI_ISL_1589882, EPI_ISL_1589883, EPI_ISL_1589884, EPI_ISL_1589885, EPI_ISL_1589886, EPI_ISL_1589887, EPI_ISL_1589888, EPI_ISL_1589889, EPI_ISL_1589890, EPI_ISL_1589891, EPI_ISL_1589892, EPI_ISL_1589893, EPI_ISL_1589894, EPI_ISL_1589895, EPI_ISL_1589896, EPI_ISL_1589897, EPI_ISL_1589898, EPI_ISL_1589899, EPI_ISL_1589900, EPI_ISL_1589901, EPI_ISL_1589902, EPI_ISL_1589903, EPI_ISL_1589904, EPI_ISL_1589905, EPI_ISL_1589906, EPI_ISL_1589907, EPI_ISL_1589908, EPI_ISL_1589909, EPI_ISL_1589910, EPI_ISL_1589911, EPI_ISL_1589912, EPI_ISL_1589913, EPI_ISL_1589914, EPI_ISL_1589915, EPI_ISL_1589916, EPI_ISL_1589917, EPI_ISL_1589918, EPI_ISL_1589919, EPI_ISL_1589920, EPI_ISL_1589921, EPI_ISL_1589922, EPI_ISL_1589923, EPI_ISL_1589924, EPI_ISL_1589925, EPI_ISL_1589926, EPI_ISL_1589927, EPI_ISL_1589928, EPI_ISL_1589929, EPI_ISL_1589930, EPI_ISL_1589931, EPI_ISL_1589932, EPI_ISL_1589933, EPI_ISL_1589934, EPI_ISL_1589935, EPI_ISL_1589936, EPI_ISL_1589937, EPI_ISL_1589938, EPI_ISL_1589939, EPI_ISL_1589940, EPI_ISL_1589941, EPI_ISL_1589942, EPI_ISL_1589943, EPI_ISL_1589944, EPI_ISL_1589945, EPI_ISL_1589946, EPI_ISL_1589947, EPI_ISL_1589948, EPI_ISL_1589949, EPI_ISL_1589950, EPI_ISL_1589951, EPI_ISL_1589952, EPI_ISL_1589953, EPI_ISL_1589954, EPI_ISL_1589955, EPI_ISL_1589956, EPI_ISL_1589957, EPI_ISL_1589958, EPI_ISL_1589959, EPI_ISL_1589960, EPI_ISL_1589961, EPI_ISL_1589962, EPI_ISL_1589963, EPI_ISL_1589964, EPI_ISL_1589965, EPI_ISL_1589966, EPI_ISL_1589967, EPI_ISL_1589968, EPI_ISL_1589973, EPI_ISL_1589974, EPI_ISL_1589975, EPI_ISL_1589976 | | | |
| see above | INSACOG-WB | National Institute of Biomedical Genomics - INSACOG | Arindam Maitra, Bhaswati Bandyopadhyay, Nidhan Kumar Biswas, Tamal Ghosh, Sreedhar Chinnaswamy, Ajay Chakraborti, Saumitra Das |
| EPI_ISL_1595862 | BBMP Urban PHC | INSACOG-KA, NIMHANS | Chitra Pattabiraman, Pramada Prasad, Anson Kunjumon George, Ananthapadmanabha Kotambail, Darshan Sreenivas, Chetan G K, Gautham Arunachal Udupi, Anita S Desai, V Ravi |
| EPI_ISL_1595866 | BANGALORE MEDICAL COLLEGE AND RESEARCH INSTITUTE | INSACOG-KA, NIMHANS | Chitra Pattabiraman, Pramada Prasad, Anson Kunjumon George, Ananthapadmanabha Kotambail, Darshan Sreenivas, Chetan G K, Gautham Arunachal Udupi, Anita S Desai, V Ravi |
| EPI_ISL_1595882 | WENLOCK HOSPITAL, MANGALORE | INSACOG-KA, NIMHANS | Chitra Pattabiraman, Pramada Prasad, Anson Kunjumon George, Ananthapadmanabha Kotambail, Darshan Sreenivas, Chetan G K, Gautham Arunachal Udupi, Anita S Desai, V Ravi |
| EPI_ISL_1595896, EPI_ISL_1595897, EPI_ISL_1595899 | KEMPEGOWDA INTERNATIONAL AIRPORT | INSACOG-KA, NIMHANS | Chitra Pattabiraman, Pramada Prasad, Anson Kunjumon George, Ananthapadmanabha Kotambail, Darshan Sreenivas, Chetan G K, Gautham Arunachal Udupi, Anita S Desai, V Ravi |
| EPI_ISL_1595907, EPI_ISL_1595909, EPI_ISL_1595910, EPI_ISL_1595916 | BBMP Urban PHC | INSACOG-KA, NIMHANS | Chitra Pattabiraman, Pramada Prasad, Anson Kunjumon George, Ananthapadmanabha Kotambail, Darshan Sreenivas, Chetan G K, Gautham Arunachal Udupi, Anita S Desai, V Ravi |
| EPI_ISL_1620132, EPI_ISL_1620133, EPI_ISL_1620134, EPI_ISL_1620136, EPI_ISL_1620137, EPI_ISL_1620138, EPI_ISL_1620140, EPI_ISL_1620143, EPI_ISL_1620154, EPI_ISL_1620159, EPI_ISL_1620160, EPI_ISL_1620161, EPI_ISL_1620162, EPI_ISL_1620163, EPI_ISL_1620164, EPI_ISL_1620165, EPI_ISL_1620166, EPI_ISL_1620170 | | | |
| see above | National Public Health Laboratory, National Centre for Infectious Diseases | National Public Health Laboratory, National Centre for Infectious Diseases | Tze Minn Mak, Zhenyang Zhou, Grace Jie Yin Ngan, Royce Ang, Lin Cui, Raymond Tzer Pin Lin |
| EPI_ISL_1622467 | Division of Emerging Infectious Diseases, Bureau of Infectious Diseases Diagnosis Control, Korea Disease Control and Prevention Agency | Division of Emerging Infectious Diseases, Bureau of Infectious Diseases Diagnosis Control, Korea Disease Control and Prevention Agency | Ae Kyung Park, Il-Hwan Kim, Heui Man Kim, Jeong-Min Kim, Jeong-Ah Kim, Chae Young Lee, Jin Sun No, Eun-Jin Kim |
| EPI_ISL_1634423, EPI_ISL_1634424, EPI_ISL_1634425, EPI_ISL_1634426, EPI_ISL_1634427, EPI_ISL_1634431 | National Public Health Laboratory, National Centre for Infectious Diseases | National Public Health Laboratory, National Centre for Infectious Diseases | Tze Minn Mak, Zhenyang Zhou, Grace Jie Yin Ngan, Royce Ang, Lin Cui, Raymond Tzer Pin Lin |
| EPI_ISL_1647348, EPI_ISL_1647349, EPI_ISL_1647350, EPI_ISL_1647351, EPI_ISL_1647352 | Division of Emerging Infectious Diseases, Bureau of Infectious Diseases Diagnosis Control, Korea Disease Control and Prevention Agency | Division of Emerging Infectious Diseases, Bureau of Infectious Diseases Diagnosis Control, Korea Disease Control and Prevention Agency | Ae Kyung Park, Il-Hwan Kim, Heui Man Kim, Jeong-Min Kim, Jeong-Ah Kim, Chae Young Lee, Jin Sun No, Eun-Jin Kim |
| EPI_ISL_1652097, EPI_ISL_1652098, EPI_ISL_1652101, EPI_ISL_1652102, EPI_ISL_1652105, EPI_ISL_1652106, EPI_ISL_1652109, EPI_ISL_1652118, EPI_ISL_1652119, EPI_ISL_1652122, EPI_ISL_1652532 | | | |
| see above | National Public Health Laboratory, National Centre for Infectious Diseases | National Public Health Laboratory, National Centre for Infectious Diseases | Tze Minn Mak, Zhenyang Zhou, Grace Jie Yin Ngan, Royce Ang, Lin Cui, Raymond Tzer Pin Lin |
| EPI_ISL_1662269 | REGIONAL VRDL, ICMR-RMRC BBSR | Institute of Life Sciences - INSACOG | Sunil K. Raghav, Safal Walia, Arup Ghosh, Atimukta Jha, Amol M. Kanampalliwar, Shifu Aggarwal, Rupesh Dash, Rajeeb Swain, Punit Prasad, INSACOG Consortium, Ajay Parida |
| EPI_ISL_1662270, EPI_ISL_1662271 | Molecular Laboratory, Vikash Multispeciality Hospital, Bargarh | Institute of Life Sciences - INSACOG | Sunil K. Raghav, Safal Walia, Arup Ghosh, Atimukta Jha, Amol M. Kanampalliwar, Shifu Aggarwal, Rupesh Dash, Rajeeb Swain, Punit Prasad, INSACOG Consortium, Ajay Parida |
| EPI_ISL_1662277, EPI_ISL_1662278, EPI_ISL_1662279, EPI_ISL_1662280, EPI_ISL_1662281 | Nalanda Medical College & Hospital, Patna | Institute of Life Sciences - INSACOG | Sunil K. Raghav, Safal Walia, Arup Ghosh, Atimukta Jha, Amol M. Kanampalliwar, Shifu Aggarwal, Rupesh Dash, Rajeeb Swain, Punit Prasad, INSACOG Consortium, Ajay Parida |
| EPI_ISL_1662282, EPI_ISL_1662283, EPI_ISL_1662284, EPI_ISL_1662285, EPI_ISL_1662286 | Veer Surendra Sai Institute of Medical Sciences and Research, Burla, Sambalpur | Institute of Life Sciences - INSACOG | Sunil K. Raghav, Safal Walia, Arup Ghosh, Atimukta Jha, Amol M. Kanampalliwar, Shifu Aggarwal, Rupesh Dash, Rajeeb Swain, Punit Prasad, INSACOG Consortium, Ajay Parida |
| EPI_ISL_1662288, EPI_ISL_1662289, EPI_ISL_1662291 | Dept. Of Microbiology, Lt.. Baliram Kashyap Memorial Govt. Medical college, Dimrapal, Jagdalpur | Institute of Life Sciences - INSACOG | Sunil K. Raghav, Safal Walia, Arup Ghosh, Atimukta Jha, Amol M. Kanampalliwar, Shifu Aggarwal, Rupesh Dash, Rajeeb Swain, Punit Prasad, INSACOG Consortium, Ajay Parida |
| EPI_ISL_1662293, EPI_ISL_1662294, EPI_ISL_1662295, EPI_ISL_1662296, EPI_ISL_1662297, EPI_ISL_1662298, EPI_ISL_1662300, EPI_ISL_1662301, EPI_ISL_1662303, EPI_ISL_1662304, EPI_ISL_1662305, EPI_ISL_1662306, EPI_ISL_1662307, EPI_ISL_1662308, EPI_ISL_1662310, EPI_ISL_1662311, EPI_ISL_1662312, EPI_ISL_1662313, EPI_ISL_1662314, EPI_ISL_1662315, EPI_ISL_1662317, EPI_ISL_1662319, EPI_ISL_1662320, EPI_ISL_1662323, EPI_ISL_1662324, EPI_ISL_1662325, EPI_ISL_1662326, EPI_ISL_1662327, EPI_ISL_1662329, EPI_ISL_1662330, EPI_ISL_1662331, EPI_ISL_1662332, EPI_ISL_1662333, EPI_ISL_1662334, EPI_ISL_1662335, EPI_ISL_1662336, EPI_ISL_1662338, EPI_ISL_1662339, EPI_ISL_1662340, EPI_ISL_1662341, EPI_ISL_1662342 | | | |
| see above | Pt. Jawahar Lal Nehru Memorial Medical College, Raipur, Chhattisgarh | Institute of Life Sciences - INSACOG | Sunil K. Raghav, Safal Walia, Arup Ghosh, Atimukta Jha, Amol M. Kanampalliwar, Shifu Aggarwal, Rupesh Dash, Rajeeb Swain, Punit Prasad, INSACOG Consortium, Ajay Parida |
| EPI_ISL_1662359, EPI_ISL_1662360, EPI_ISL_1662362, EPI_ISL_1662363, EPI_ISL_1662368, EPI_ISL_1662370, EPI_ISL_1662372, EPI_ISL_1662373, EPI_ISL_1662375, EPI_ISL_1662378, EPI_ISL_1662380, EPI_ISL_1662382 | | | |
| see above | Govt. Medical College, Ambikapur, Surguja | Institute of Life Sciences - INSACOG | Sunil K. Raghav, Safal Walia, Arup Ghosh, Atimukta Jha, Amol M. Kanampalliwar, Shifu Aggarwal, Rupesh Dash, Rajeeb Swain, Punit Prasad, INSACOG Consortium, Ajay Parida |
| EPI_ISL_1662391, EPI_ISL_1662393, EPI_ISL_1662394, EPI_ISL_1662395, EPI_ISL_1662396, EPI_ISL_1662398, EPI_ISL_1662400, EPI_ISL_1662406, EPI_ISL_1662407, EPI_ISL_1662408, EPI_ISL_1662410, EPI_ISL_1662411, EPI_ISL_1662412, EPI_ISL_1662414, EPI_ISL_1662415, EPI_ISL_1662416, EPI_ISL_1662417, EPI_ISL_1662418, EPI_ISL_1662420 | | | |
| see above | BRLSABVM Government Medical College, Rajnandgaon, | Institute of Life Sciences - INSACOG | Sunil K. Raghav, Safal Walia, Arup Ghosh, Atimukta Jha, Amol M. Kanampalliwar, Shifu Aggarwal, Rupesh Dash, Rajeeb Swain, Punit Prasad, INSACOG |

| | | | |
|---|---|---|---|
| | Chhattisgarh | | Consortium, Ajay Parida |
| EPI_ISL_1662427, EPI_ISL_1662431, EPI_ISL_1662434, EPI_ISL_1662439, EPI_ISL_1662442, EPI_ISL_1662443, EPI_ISL_1662444 | Capital hospital, Bhubaneswar | Institute of Life Sciences - INSACOG | Sunil K. Raghav, Safal Walia, Arup Ghosh, Atimukta Jha, Amol M. Kanampalliwar, Shifu Aggarwal, Rupesh Dash, Rajeeb Swain, Punit Prasad, INSACOG Consortium, Ajay Parida |
| EPI_ISL_1662447, EPI_ISL_1662450, EPI_ISL_1662451, EPI_ISL_1662452, EPI_ISL_1662462 | Immunogenomics lab, Institute of Life Sciences, Bhubaneswar | Institute of Life Sciences - INSACOG | Sunil K. Raghav, Safal Walia, Arup Ghosh, Atimukta Jha, Amol M. Kanampalliwar, Shifu Aggarwal, Rupesh Dash, Rajeeb Swain, Punit Prasad, INSACOG Consortium, Ajay Parida |
| EPI_ISL_1663235, EPI_ISL_1663236, EPI_ISL_1663238, EPI_ISL_1663239, EPI_ISL_1663240, EPI_ISL_1663241, EPI_ISL_1663243, EPI_ISL_1663246, EPI_ISL_1663247, EPI_ISL_1663250, EPI_ISL_1663251 | | | |
| see above | BRLSABVM Government Medical College, Rajnandgaon, Chhattisgarh | Institute of Life Sciences - INSACOG | Sunil K. Raghav, Safal Walia, Arup Ghosh, Atimukta Jha, Amol M. Kanampalliwar, Shifu Aggarwal, Rupesh Dash, Rajeeb Swain, Punit Prasad, INSACOG Consortium, Ajay Parida |
| EPI_ISL_1663263, EPI_ISL_1663264, EPI_ISL_1663265, EPI_ISL_1663270, EPI_ISL_1663271, EPI_ISL_1663272, EPI_ISL_1663273, EPI_ISL_1663274, EPI_ISL_1663303, EPI_ISL_1663304, EPI_ISL_1663305, EPI_ISL_1663306, EPI_ISL_1663307, EPI_ISL_1663308, EPI_ISL_1663310, EPI_ISL_1663311, EPI_ISL_1663312, EPI_ISL_1663313, EPI_ISL_1663315, EPI_ISL_1663316, EPI_ISL_1663317, EPI_ISL_1663318, EPI_ISL_1663320, EPI_ISL_1663321, EPI_ISL_1663322 | | | |
| see above | CIIMS, Bilaspur, Chhattisgarh | Institute of Life Sciences - INSACOG | Sunil K. Raghav, Safal Walia, Arup Ghosh, Atimukta Jha, Amol M. Kanampalliwar, Shifu Aggarwal, Rupesh Dash, Rajeeb Swain, Punit Prasad, INSACOG Consortium, Ajay Parida |
| EPI_ISL_1663323, EPI_ISL_1663324 | Capital hospital, Bhubaneswar | Institute of Life Sciences - INSACOG | Sunil K. Raghav, Safal Walia, Arup Ghosh, Atimukta Jha, Amol M. Kanampalliwar, Shifu Aggarwal, Rupesh Dash, Rajeeb Swain, Punit Prasad, INSACOG Consortium, Ajay Parida |
| EPI_ISL_1663325, EPI_ISL_1663326, EPI_ISL_1663327, EPI_ISL_1663329, EPI_ISL_1663330, EPI_ISL_1663332 | Veer Surendra Sai Institute of Medical Sciences and Research, Burla, Sambalpur | Institute of Life Sciences - INSACOG | Sunil K. Raghav, Safal Walia, Arup Ghosh, Atimukta Jha, Amol M. Kanampalliwar, Shifu Aggarwal, Rupesh Dash, Rajeeb Swain, Punit Prasad, INSACOG Consortium, Ajay Parida |
| EPI_ISL_1663333, EPI_ISL_1663334 | PRMMCCH, Baripada | Institute of Life Sciences - INSACOG | Sunil K. Raghav, Safal Walia, Arup Ghosh, Atimukta Jha, Amol M. Kanampalliwar, Shifu Aggarwal, Rupesh Dash, Rajeeb Swain, Punit Prasad, INSACOG Consortium, Ajay Parida |
| EPI_ISL_1663338, EPI_ISL_1663339, EPI_ISL_1663340, EPI_ISL_1663343, EPI_ISL_1663344, EPI_ISL_1663347, EPI_ISL_1663350, EPI_ISL_1663352 | RIMS, Ranchi | Institute of Life Sciences - INSACOG | Sunil K. Raghav, Safal Walia, Arup Ghosh, Atimukta Jha, Amol M. Kanampalliwar, Shifu Aggarwal, Rupesh Dash, Rajeeb Swain, Punit Prasad, INSACOG Consortium, Ajay Parida |
| EPI_ISL_1663354 | ITKI Arogyashala | Institute of Life Sciences - INSACOG | Sunil K. Raghav, Safal Walia, Arup Ghosh, Atimukta Jha, Amol M. Kanampalliwar, Shifu Aggarwal, Rupesh Dash, Rajeeb Swain, Punit Prasad, INSACOG Consortium, Ajay Parida |
| EPI_ISL_1663357 | Capital hospital, Bhubaneswar | Institute of Life Sciences - INSACOG | Sunil K. Raghav, Safal Walia, Arup Ghosh, Atimukta Jha, Amol M. Kanampalliwar, Shifu Aggarwal, Rupesh Dash, Rajeeb Swain, Punit Prasad, INSACOG Consortium, Ajay Parida |
| EPI_ISL_1663358, EPI_ISL_1663362 | RMRI, Patna | Institute of Life Sciences - INSACOG | Sunil K. Raghav, Safal Walia, Arup Ghosh, Atimukta Jha, Amol M. Kanampalliwar, Shifu Aggarwal, Rupesh Dash, Rajeeb Swain, Punit Prasad, INSACOG Consortium, Ajay Parida |
| EPI_ISL_1663363, EPI_ISL_1663365, EPI_ISL_1663366, EPI_ISL_1663367, EPI_ISL_1663368, EPI_ISL_1663369, EPI_ISL_1663370, EPI_ISL_1663375, EPI_ISL_1663376, EPI_ISL_1663377, EPI_ISL_1663378, EPI_ISL_1663379, EPI_ISL_1663380, EPI_ISL_1663381, EPI_ISL_1663382, EPI_ISL_1663383, EPI_ISL_1663384, EPI_ISL_1663385, EPI_ISL_1663386, EPI_ISL_1663387, EPI_ISL_1663388, EPI_ISL_1663389, EPI_ISL_1663391, EPI_ISL_1663393, EPI_ISL_1663395, EPI_ISL_1663397, EPI_ISL_1663398, EPI_ISL_1663400, EPI_ISL_1663401, EPI_ISL_1663403, EPI_ISL_1663405, EPI_ISL_1663406, EPI_ISL_1663407, EPI_ISL_1663408, EPI_ISL_1663409, EPI_ISL_1663410, EPI_ISL_1663411, EPI_ISL_1663415, EPI_ISL_1663416, EPI_ISL_1663417, EPI_ISL_1663418, EPI_ISL_1663419, EPI_ISL_1663420, EPI_ISL_1663421, EPI_ISL_1663422, EPI_ISL_1663423, EPI_ISL_1663424, EPI_ISL_1663426, EPI_ISL_1663429, EPI_ISL_1663433, EPI_ISL_1663434, EPI_ISL_1663435, EPI_ISL_1663437, EPI_ISL_1663443, EPI_ISL_1663444, EPI_ISL_1663448, EPI_ISL_1663452, EPI_ISL_1663454, EPI_ISL_1663455, EPI_ISL_1663456, EPI_ISL_1663458, EPI_ISL_1663459, EPI_ISL_1663460, EPI_ISL_1663463, EPI_ISL_1663466, EPI_ISL_1663469, EPI_ISL_1663471, EPI_ISL_1663472, EPI_ISL_1663474, EPI_ISL_1663476, EPI_ISL_1663481, EPI_ISL_1663487 | | | |
| see above | NCCS, Pune | Institute of Life Sciences - INSACOG | Sunil K. Raghav, Safal Walia, Arup Ghosh, Atimukta Jha, Amol M. Kanampalliwar, Shifu Aggarwal, Rupesh Dash, Rajeeb Swain, Punit Prasad, INSACOG Consortium, Ajay Parida |
| EPI_ISL_1663489, EPI_ISL_1663490, EPI_ISL_1663491, EPI_ISL_1663493, EPI_ISL_1663494, EPI_ISL_1663495, EPI_ISL_1663496 | Capital hospital, Bhubaneswar | Institute of Life Sciences - INSACOG | Sunil K. Raghav, Safal Walia, Arup Ghosh, Atimukta Jha, Amol M. Kanampalliwar, Shifu Aggarwal, Rupesh Dash, Rajeeb Swain, Punit Prasad, INSACOG Consortium, Ajay Parida |
| EPI_ISL_1663497, EPI_ISL_1663498, EPI_ISL_1663499, EPI_ISL_1663500, EPI_ISL_1663501, EPI_ISL_1663502, EPI_ISL_1663503 | Veer Surendra Sai Institute of Medical Sciences and Research, Burla, Sambalpur | Institute of Life Sciences - INSACOG | Sunil K. Raghav, Safal Walia, Arup Ghosh, Atimukta Jha, Amol M. Kanampalliwar, Shifu Aggarwal, Rupesh Dash, Rajeeb Swain, Punit Prasad, INSACOG Consortium, Ajay Parida |
| EPI_ISL_1663504, EPI_ISL_1663507, EPI_ISL_1663509, EPI_ISL_1663514, EPI_ISL_1663518, EPI_ISL_1663519, EPI_ISL_1663521, EPI_ISL_1663523 | AIIMS, Patna | Institute of Life Sciences - INSACOG | Sunil K. Raghav, Safal Walia, Arup Ghosh, Atimukta Jha, Amol M. Kanampalliwar, Shifu Aggarwal, Rupesh Dash, Rajeeb Swain, Punit Prasad, INSACOG Consortium, Ajay Parida |
| EPI_ISL_1663524, EPI_ISL_1663525, EPI_ISL_1663526, EPI_ISL_1663527, EPI_ISL_1663529, EPI_ISL_1663530, EPI_ISL_1663531, EPI_ISL_1663535 | REGIONAL VRDL, ICMR-RMRC BBSR | Institute of Life Sciences - INSACOG | Sunil K. Raghav, Safal Walia, Arup Ghosh, Atimukta Jha, Amol M. Kanampalliwar, Shifu Aggarwal, Rupesh Dash, Rajeeb Swain, Punit Prasad, INSACOG Consortium, Ajay Parida |
| EPI_ISL_1663539, EPI_ISL_1663540, EPI_ISL_1663541, EPI_ISL_1663545, EPI_ISL_1663546, EPI_ISL_1663548 | MGM Medical College, Jamshedpur | Institute of Life Sciences - INSACOG | Sunil K. Raghav, Safal Walia, Arup Ghosh, Atimukta Jha, Amol M. Kanampalliwar, Shifu Aggarwal, Rupesh Dash, Rajeeb Swain, Punit Prasad, INSACOG Consortium, Ajay Parida |
| EPI_ISL_1663552, EPI_ISL_1663553, EPI_ISL_1663554, EPI_ISL_1663555, EPI_ISL_1663557, EPI_ISL_1663558, EPI_ISL_1663559, EPI_ISL_1663560, EPI_ISL_1663561, EPI_ISL_1663562, EPI_ISL_1663563, EPI_ISL_1663564 | | | |
| see above | Immunogenomics lab, Institute of Life Sciences, Bhubaneswar | Institute of Life Sciences - INSACOG | Sunil K. Raghav, Safal Walia, Arup Ghosh, Atimukta Jha, Amol M. Kanampalliwar, Shifu Aggarwal, Rupesh Dash, Rajeeb Swain, Punit Prasad, INSACOG Consortium, Ajay Parida |
| EPI_ISL_1677768 | Gujarat Biotechnology Research Centre (GBRC), Gandhinagar | Gujarat Biotechnology Research Centre | Nitin Savaliya,Dinesh Kumar,Twinkle Soni,Sonal Sharma ,Zuber Saiyed,Ramesh Pandit,Janvi Raval,Zarna Patel,Umang Mishra,Nitesh Shah,Chaitanya Joshi,Madhvi Joshi |
| EPI_ISL_1703892, EPI_ISL_1703898, EPI_ISL_1703901, EPI_ISL_1703906, EPI_ISL_1703908, EPI_ISL_1703909, EPI_ISL_1703913, EPI_ISL_1703914, EPI_ISL_1703915, EPI_ISL_1703916, EPI_ISL_1703917, EPI_ISL_1703918, EPI_ISL_1703922, EPI_ISL_1703923, EPI_ISL_1703924, EPI_ISL_1703927, EPI_ISL_1703928, EPI_ISL_1703929, EPI_ISL_1703930, EPI_ISL_1703931, EPI_ISL_1703932, EPI_ISL_1703933, EPI_ISL_1703934, EPI_ISL_1703935, EPI_ISL_1703937, EPI_ISL_1703938, EPI_ISL_1703939, EPI_ISL_1703940, EPI_ISL_1703941, EPI_ISL_1703942, EPI_ISL_1703943, EPI_ISL_1703945, EPI_ISL_1703946, EPI_ISL_1703948, EPI_ISL_1703949, EPI_ISL_1703950, EPI_ISL_1703951, EPI_ISL_1703952, EPI_ISL_1703953, EPI_ISL_1703954, EPI_ISL_1703955, EPI_ISL_1703956, EPI_ISL_1703957, EPI_ISL_1703959, EPI_ISL_1703960, EPI_ISL_1703961, EPI_ISL_1703963, EPI_ISL_1703964, EPI_ISL_1703971, EPI_ISL_1703972, EPI_ISL_1703973, EPI_ISL_1703974, EPI_ISL_1703975, EPI_ISL_1703982, EPI_ISL_1703983, EPI_ISL_1703984, EPI_ISL_1703986, EPI_ISL_1703989, EPI_ISL_1703990, EPI_ISL_1703992, EPI_ISL_1703993, EPI_ISL_1703994, EPI_ISL_1703996, EPI_ISL_1704008, EPI_ISL_1704009, EPI_ISL_1704014, EPI_ISL_1704015, EPI_ISL_1704016, EPI_ISL_1704017, EPI_ISL_1704018, EPI_ISL_1704019, EPI_ISL_1704020, EPI_ISL_1704021, EPI_ISL_1704022, EPI_ISL_1704024, EPI_ISL_1704025, EPI_ISL_1704026, EPI_ISL_1704027, EPI_ISL_1704028, EPI_ISL_1704029, EPI_ISL_1704030, EPI_ISL_1704032, EPI_ISL_1704033, EPI_ISL_1704034, EPI_ISL_1704035, EPI_ISL_1704036, EPI_ISL_1704037, EPI_ISL_1704042, EPI_ISL_1704043, EPI_ISL_1704093, EPI_ISL_1704094, EPI_ISL_1704097, EPI_ISL_1704099, EPI_ISL_1704100, EPI_ISL_1704101, EPI_ISL_1704102, EPI_ISL_1704110, EPI_ISL_1704111, EPI_ISL_1704113, EPI_ISL_1704114, EPI_ISL_1704115, EPI_ISL_1704116, EPI_ISL_1704117, EPI_ISL_1704118, EPI_ISL_1704119, EPI_ISL_1704127, EPI_ISL_1704128, EPI_ISL_1704149, EPI_ISL_1704150, EPI_ISL_1704151, EPI_ISL_1704152, EPI_ISL_1704153, EPI_ISL_1704154, EPI_ISL_1704158, EPI_ISL_1704159, EPI_ISL_1704161, EPI_ISL_1704196, EPI_ISL_1704197, EPI_ISL_1704198, EPI_ISL_1704199, EPI_ISL_1704200, EPI_ISL_1704201, EPI_ISL_1704202, EPI_ISL_1704227, EPI_ISL_1704229, EPI_ISL_1704230, EPI_ISL_1704231, EPI_ISL_1704232, EPI_ISL_1704233, EPI_ISL_1704234, EPI_ISL_1704235, EPI_ISL_1704236, EPI_ISL_1704237, EPI_ISL_1704238, EPI_ISL_1704240, EPI_ISL_1704241, EPI_ISL_1704242, EPI_ISL_1704243, EPI_ISL_1704244, EPI_ISL_1704245, EPI_ISL_1704246, EPI_ISL_1704247, EPI_ISL_1704249, EPI_ISL_1704250, EPI_ISL_1704277, EPI_ISL_1704335, EPI_ISL_1704336, EPI_ISL_1704337, EPI_ISL_1704338, EPI_ISL_1704339, EPI_ISL_1704340, EPI_ISL_1704341, EPI_ISL_1704342, EPI_ISL_1704343, EPI_ISL_1704344, EPI_ISL_1704345, EPI_ISL_1704346, EPI_ISL_1704347, EPI_ISL_1704348, EPI_ISL_1704349, EPI_ISL_1704350, EPI_ISL_1704355, EPI_ISL_1704356, EPI_ISL_1704357, EPI_ISL_1704358, EPI_ISL_1704359, EPI_ISL_1704360, EPI_ISL_1704361, EPI_ISL_1704364, EPI_ISL_1704365, EPI_ISL_1704367, EPI_ISL_1704368, EPI_ISL_1704369, EPI_ISL_1704370, EPI_ISL_1704372, EPI_ISL_1704373, EPI_ISL_1704374, EPI_ISL_1704375, EPI_ISL_1704377, EPI_ISL_1704378, EPI_ISL_1704379, EPI_ISL_1704380, EPI_ISL_1704394, EPI_ISL_1704395, EPI_ISL_1704396, EPI_ISL_1704397, EPI_ISL_1704398, EPI_ISL_1704399, EPI_ISL_1704400, EPI_ISL_1704401, EPI_ISL_1704402, EPI_ISL_1704404, EPI_ISL_1704405, EPI_ISL_1704406, EPI_ISL_1704407, EPI_ISL_1704408, EPI_ISL_1704410, EPI_ISL_1704411, EPI_ISL_1704412, EPI_ISL_1704413, EPI_ISL_1704414, EPI_ISL_1704415, EPI_ISL_1704416, EPI_ISL_1704417, | | | |

| | | | |
|---|---|---|---|
| EPI_ISL_1704418, EPI_ISL_1704419, EPI_ISL_1704420, EPI_ISL_1704421, EPI_ISL_1704422, EPI_ISL_1704423, EPI_ISL_1704424, EPI_ISL_1704425, EPI_ISL_1704426, EPI_ISL_1704427, EPI_ISL_1704428, EPI_ISL_1704429, EPI_ISL_1704430, EPI_ISL_1704431, EPI_ISL_1704432, EPI_ISL_1704433, EPI_ISL_1704434, EPI_ISL_1704435, EPI_ISL_1704436, EPI_ISL_1704437, EPI_ISL_1704438, EPI_ISL_1704439, EPI_ISL_1704440, EPI_ISL_1704441, EPI_ISL_1704442, EPI_ISL_1704443, EPI_ISL_1704444, EPI_ISL_1704445, EPI_ISL_1704446, EPI_ISL_1704447, EPI_ISL_1704448, EPI_ISL_1704449, EPI_ISL_1704450, EPI_ISL_1704451, EPI_ISL_1704452, EPI_ISL_1704453, EPI_ISL_1704454, EPI_ISL_1704455, EPI_ISL_1704456, EPI_ISL_1704457, EPI_ISL_1704458, EPI_ISL_1704459, EPI_ISL_1704460, EPI_ISL_1704461, EPI_ISL_1704462, EPI_ISL_1704463, EPI_ISL_1704464, EPI_ISL_1704465, EPI_ISL_1704466, EPI_ISL_1704467, EPI_ISL_1704468, EPI_ISL_1704469, EPI_ISL_1704470, EPI_ISL_1704471, EPI_ISL_1704472, EPI_ISL_1704473, EPI_ISL_1704474, EPI_ISL_1704475, EPI_ISL_1704476, EPI_ISL_1704477, EPI_ISL_1704478, EPI_ISL_1704479, EPI_ISL_1704480, EPI_ISL_1704481, EPI_ISL_1704482, EPI_ISL_1704483, EPI_ISL_1704484, EPI_ISL_1704485, EPI_ISL_1704486, EPI_ISL_1704487, EPI_ISL_1704488, EPI_ISL_1704489, EPI_ISL_1704490, EPI_ISL_1704491, EPI_ISL_1704492, EPI_ISL_1704494, EPI_ISL_1704495, EPI_ISL_1704496, EPI_ISL_1704497, EPI_ISL_1704498, EPI_ISL_1704499, EPI_ISL_1704500, EPI_ISL_1704501, EPI_ISL_1704502, EPI_ISL_1704503, EPI_ISL_1704504, EPI_ISL_1704505, EPI_ISL_1704506, EPI_ISL_1704507, EPI_ISL_1704508, EPI_ISL_1704509, EPI_ISL_1704510, EPI_ISL_1704511, EPI_ISL_1704512, EPI_ISL_1704513, EPI_ISL_1704514, EPI_ISL_1704515, EPI_ISL_1704516, EPI_ISL_1704517, EPI_ISL_1704518, EPI_ISL_1704519, EPI_ISL_1704520, EPI_ISL_1704521, EPI_ISL_1704522, EPI_ISL_1704523, EPI_ISL_1704524, EPI_ISL_1704525, EPI_ISL_1704526, EPI_ISL_1704527, EPI_ISL_1704528, EPI_ISL_1704529, EPI_ISL_1704530, EPI_ISL_1704531, EPI_ISL_1704532, EPI_ISL_1704533, EPI_ISL_1704534, EPI_ISL_1704535, EPI_ISL_1704536, EPI_ISL_1704537, EPI_ISL_1704538, EPI_ISL_1704539, EPI_ISL_1704540, EPI_ISL_1704541, EPI_ISL_1704542, EPI_ISL_1704543, EPI_ISL_1704544, EPI_ISL_1704545, EPI_ISL_1704546, EPI_ISL_1704547, EPI_ISL_1704548, EPI_ISL_1704549, EPI_ISL_1704550, EPI_ISL_1704551, EPI_ISL_1704552, EPI_ISL_1704553, EPI_ISL_1704554, EPI_ISL_1704555, EPI_ISL_1704556, EPI_ISL_1704557, EPI_ISL_1704558, EPI_ISL_1704559, EPI_ISL_1704560, EPI_ISL_1704561, EPI_ISL_1704562, EPI_ISL_1704563, EPI_ISL_1704564, EPI_ISL_1704565, EPI_ISL_1704566, EPI_ISL_1704567, EPI_ISL_1704568, EPI_ISL_1704569, EPI_ISL_1704570, EPI_ISL_1704571, EPI_ISL_1704572, EPI_ISL_1704573, EPI_ISL_1704574, EPI_ISL_1704575, EPI_ISL_1704576, EPI_ISL_1704577, EPI_ISL_1704578, EPI_ISL_1704579, EPI_ISL_1704580, EPI_ISL_1704581, EPI_ISL_1704585, EPI_ISL_1704586, EPI_ISL_1704587, EPI_ISL_1704588, EPI_ISL_1704589, EPI_ISL_1704590, EPI_ISL_1704591, EPI_ISL_1704592, EPI_ISL_1704593, EPI_ISL_1704594, EPI_ISL_1704595, EPI_ISL_1704596, EPI_ISL_1704597, EPI_ISL_1704598, EPI_ISL_1704599, EPI_ISL_1704600, EPI_ISL_1704601, EPI_ISL_1704602, EPI_ISL_1704603, EPI_ISL_1704604, EPI_ISL_1704605, EPI_ISL_1704606, EPI_ISL_1704607, EPI_ISL_1704608, EPI_ISL_1704609, EPI_ISL_1704610, EPI_ISL_1704611, EPI_ISL_1704612, EPI_ISL_1704613, EPI_ISL_1704614, EPI_ISL_1704615, EPI_ISL_1704618, EPI_ISL_1704619, EPI_ISL_1704621, EPI_ISL_1704622, EPI_ISL_1704623, EPI_ISL_1704624, EPI_ISL_1704625, EPI_ISL_1704626, EPI_ISL_1704627, EPI_ISL_1704628, EPI_ISL_1704629, EPI_ISL_1704630, EPI_ISL_1704631, EPI_ISL_1704632, EPI_ISL_1704633, EPI_ISL_1704634, EPI_ISL_1704635, EPI_ISL_1704636, EPI_ISL_1704637, EPI_ISL_1704638, EPI_ISL_1704639, EPI_ISL_1704663 | | | |
| see above | ICMR-National Institute of Virology - INSACOG | NIV Influenza | Dr. Varsha Potdar |
| EPI_ISL_1704810, EPI_ISL_1704811, EPI_ISL_1704813, EPI_ISL_1704814, EPI_ISL_1704815, EPI_ISL_1704818, EPI_ISL_1704823, EPI_ISL_1704825, EPI_ISL_1704826, EPI_ISL_1704827, EPI_ISL_1704828, EPI_ISL_1704830, EPI_ISL_1704831, EPI_ISL_1704832, EPI_ISL_1704833, EPI_ISL_1704834, EPI_ISL_1704835, EPI_ISL_1704836, EPI_ISL_1704839, EPI_ISL_1704840, EPI_ISL_1704841, EPI_ISL_1704842, EPI_ISL_1704844 | | | |
| see above | National Public Health Laboratory, National Centre for Infectious Diseases | National Public Health Laboratory, National Centre for Infectious Diseases | Tze Minn Mak, Zhenyang Zhou, Grace Jie Yin Ngan, Royce Ang, Lin Cui, Raymond Tzer Pin Lin |
| EPI_ISL_1708470, EPI_ISL_1708473 | Dr.S.raju Director of public heath and preventive medicine | inStem NCBS - INSACOG | Uma Ramakrishnan Dasaradhi Palakodeti Aswin SaiNarain |
| EPI_ISL_1708474, EPI_ISL_1708475, EPI_ISL_1708476, EPI_ISL_1708477, EPI_ISL_1708478, EPI_ISL_1708479, EPI_ISL_1708483, EPI_ISL_1708484, EPI_ISL_1708485, EPI_ISL_1708486, EPI_ISL_1708487, EPI_ISL_1708488, EPI_ISL_1708489, EPI_ISL_1708490, EPI_ISL_1708491, EPI_ISL_1708492, EPI_ISL_1708493, EPI_ISL_1708494, EPI_ISL_1708495, EPI_ISL_1708496, EPI_ISL_1708497 | | | |
| see above | Dr.Madhusudhan MBBD MD ICMR-VRDL | inStem NCBS - INSACOG | Uma Ramakrishnan Dasaradhi Palakodeti Aswin SaiNarain |
| EPI_ISL_1708498, EPI_ISL_1708501, EPI_ISL_1708502, EPI_ISL_1708503 | Dr.S.raju Director of public heath and preventive medicine | inStem NCBS - INSACOG | Aswin Seshasayee" |
| EPI_ISL_1710598 | Communicable Diseases, Interactive Research School for Health Affairs | Communicable Diseases, Interactive Research School for Health Affairs | Shubham Shrivastava, Meera Modak, Rashmi Virkar, Shamburaje S Pisal, Akhilesh Chandra Mishra, Vidya A Arankalle |
| EPI_ISL_1719868, EPI_ISL_1719870, EPI_ISL_1719873, EPI_ISL_1719878, EPI_ISL_1719879, EPI_ISL_1719881, EPI_ISL_1719882, EPI_ISL_1719884, EPI_ISL_1719885, EPI_ISL_1719887, EPI_ISL_1719889, EPI_ISL_1719891, EPI_ISL_1719892, EPI_ISL_1719895, EPI_ISL_1719896, EPI_ISL_1719897, EPI_ISL_1719898, EPI_ISL_1719899 | | | |
| see above | National Public Health Laboratory, National Centre for Infectious Diseases | National Public Health Laboratory, National Centre for Infectious Diseases | Tze Minn Mak, Zhenyang Zhou, Grace Jie Yin Ngan, Royce Ang, Lin Cui, Raymond Tzer Pin Lin |
| EPI_ISL_1731751, EPI_ISL_1731756 | State Virus Research and Diagnostic Laboratory (VRDL), AIIMS Raipur | State Virus Research and Diagnostic Laboratory (VRDL), AIIMS Raipur | Pushpendra Singh, Kuldeep Sharma, Priyanka Singh, Somya Sharma, Sanjay Singh Negi, Anudita Bhargava |
| EPI_ISL_1745236 | ULSS 7 Pedemontana - Distretto 2 | Istituto Zooprofilattico Sperimentale delle Venezie | Adelaide Milani, Alessia Schivo, Annalisa Salviato, Elisa Palumbo, Erika Giorgia Quaranta, Luca Tassoni, Ambra Pastori, Edoardo Giussani, Alice Fusaro, Isabella Monne, Calogero Terregino, Antonia Ricci |
| EPI_ISL_1752677 | National Public Health Laboratory, National Centre for Infectious Diseases | National Public Health Laboratory, National Centre for Infectious Diseases | Tze Minn Mak, Zhenyang Zhou, Grace Jie Yin Ngan, Royce Ang, Lin Cui, Raymond Tzer Pin Lin |
| EPI_ISL_1789542 | WHO National Influenza Centre Russian Federation | WHO National Influenza Centre Russian Federation | Andrey Komissarov, Artem Fadeev, Kseniya Komissarova, Oula Mansour, Mikhail Bakaev, Tamila Musaeva, Maria Timofeeva, Veronika Eder, Maria Pisareva, Daria Danilenko, Ksenia Safina, Elena Nabieva, Georgii Bazykin, Dmitry Lioznov |
| EPI_ISL_1816925, EPI_ISL_1816926, EPI_ISL_1816928, EPI_ISL_1816929, EPI_ISL_1816938, EPI_ISL_1816939, EPI_ISL_1816940, EPI_ISL_1816941, EPI_ISL_1816942, EPI_ISL_1816943, EPI_ISL_1816944, EPI_ISL_1816945, EPI_ISL_1816947, EPI_ISL_1816948, EPI_ISL_1816949, EPI_ISL_1816950, EPI_ISL_1816951, EPI_ISL_1816952, EPI_ISL_1816953, EPI_ISL_1816954, EPI_ISL_1816955, EPI_ISL_1816957, EPI_ISL_1816958, EPI_ISL_1816959, EPI_ISL_1816960, EPI_ISL_1816961, EPI_ISL_1816963, EPI_ISL_1816964, EPI_ISL_1816965, EPI_ISL_1816966, EPI_ISL_1816967, EPI_ISL_1816968, EPI_ISL_1816971, EPI_ISL_1816972, EPI_ISL_1816973 | | | |
| see above | National Public Health Laboratory, National Centre for Infectious Diseases | National Public Health Laboratory, National Centre for Infectious Diseases | Tze Minn Mak, Zhenyang Zhou, Grace Jie Yin Ngan, Royce Ang, Lin Cui, Raymond Tzer Pin Lin |
| EPI_ISL_1818586 | COMMAND HOSPITAL | INSACOG-KA, NIMHANS | Chitra Pattabiraman, Pramada Prasad, Anson Kunjumon George, Ananthapadmanabha Kotambail, Darshan Sreenivas, Chetan G K, Gautham Arunachal Udupi, Anita S Desai, V Ravi |
| EPI_ISL_1818590 | National Institute of Mental Health and Neurosciences (NIMHANS) | INSACOG-KA, NIMHANS | Chitra Pattabiraman, Pramada Prasad, Anson Kunjumon George, Ananthapadmanabha Kotambail, Darshan Sreenivas, Chetan G K, Gautham Arunachal Udupi, Anita S Desai, V Ravi |
| EPI_ISL_1818594 | RAILWAY H | INSACOG-KA, NIMHANS | Chitra Pattabiraman, Pramada Prasad, Anson Kunjumon George, Ananthapadmanabha Kotambail, Darshan Sreenivas, Chetan G K, Gautham Arunachal Udupi, Anita S Desai, V Ravi |
| EPI_ISL_1818595 | BBMP Urban PHC | INSACOG-KA, NIMHANS | Chitra Pattabiraman, Pramada Prasad, Anson Kunjumon George, Ananthapadmanabha Kotambail, Darshan Sreenivas, Chetan G K, Gautham Arunachal Udupi, Anita S Desai, V Ravi |
| EPI_ISL_1818598, EPI_ISL_1818600 | National Institute of Mental Health and Neurosciences (NIMHANS) | INSACOG-KA, NIMHANS | Chitra Pattabiraman, Pramada Prasad, Anson Kunjumon George, Ananthapadmanabha Kotambail, Darshan Sreenivas, Chetan G K, Gautham Arunachal Udupi, Anita S Desai, V Ravi |
| EPI_ISL_1818617, EPI_ISL_1818622 | KEMPEGOWDA INTERNATIONAL AIRPORT | INSACOG-KA, NIMHANS | Chitra Pattabiraman, Pramada Prasad, Anson Kunjumon George, Ananthapadmanabha Kotambail, Darshan Sreenivas, Chetan G K, Gautham Arunachal Udupi, Anita S Desai, V Ravi |
| EPI_ISL_1818630 | Jayanagar General Hospital | INSACOG-KA, NIMHANS | Chitra Pattabiraman, Pramada Prasad, Anson Kunjumon George, Ananthapadmanabha Kotambail, Darshan Sreenivas, Chetan G K, Gautham Arunachal Udupi, Anita S Desai, V Ravi |
| EPI_ISL_1818631, EPI_ISL_1818634, EPI_ISL_1818637 | BBMP Urban PHC | INSACOG-KA, NIMHANS | Chitra Pattabiraman, Pramada Prasad, Anson Kunjumon George, Ananthapadmanabha Kotambail, Darshan Sreenivas, Chetan G K, Gautham Arunachal Udupi, Anita S Desai, V Ravi |
| EPI_ISL_1824680 | Virology Laboratory, Scientific Department, Army Medical Center | Virology Laboratory, Scientific Department, Army Medical Center | Silvia Fillo, Riccardo De Sanctis, Antonella Fortunato, Anella Monte, Rossella Brandi, Giulia Campoli, Marzia Cavalli, Lucia Nicosia, Stella Lia, Anna Anselmo, Vanessa Vera Fain, Francesco Giordani, Giandomenico Cerreto, Filippo Molinari, Giancarlo Petralito, Florigio Lista |
| EPI_ISL_1837991, EPI_ISL_1837998, EPI_ISL_1838004, EPI_ISL_1838007, EPI_ISL_1838010, EPI_ISL_1838012, EPI_ISL_1838015, EPI_ISL_1838018, EPI_ISL_1838023, EPI_ISL_1838026, EPI_ISL_1838032, EPI_ISL_1838035, EPI_ISL_1838037, EPI_ISL_1838040, EPI_ISL_1838042, EPI_ISL_1838045, EPI_ISL_1838050, EPI_ISL_1838052, EPI_ISL_1838063, EPI_ISL_1838068, EPI_ISL_1838073, EPI_ISL_1838075, EPI_ISL_1838078, EPI_ISL_1838080, EPI_ISL_1838097, EPI_ISL_1838100, EPI_ISL_1838102, EPI_ISL_1838106, EPI_ISL_1838109, EPI_ISL_1838114, EPI_ISL_1838117, EPI_ISL_1838119, EPI_ISL_1838122, EPI_ISL_1838123, EPI_ISL_1838125, EPI_ISL_1838131, EPI_ISL_1838132, EPI_ISL_1838134, EPI_ISL_1838136, EPI_ISL_1838140, EPI_ISL_1838142, EPI_ISL_1838143, EPI_ISL_1838151, EPI_ISL_1838152, EPI_ISL_1838155, EPI_ISL_1838156, EPI_ISL_1838158, EPI_ISL_1838159, EPI_ISL_1838162, EPI_ISL_1838164, EPI_ISL_1838165, EPI_ISL_1838168, EPI_ISL_1838169, EPI_ISL_1838173, EPI_ISL_1838179, EPI_ISL_1838185, EPI_ISL_1838186, EPI_ISL_1838188, EPI_ISL_1838194, EPI_ISL_1838195, EPI_ISL_1838198, EPI_ISL_1838200, EPI_ISL_1838205, EPI_ISL_1838207, EPI_ISL_1838209, EPI_ISL_1838212, EPI_ISL_1838214, EPI_ISL_1838217, EPI_ISL_1838219, EPI_ISL_1838220, EPI_ISL_1838222, EPI_ISL_1838225, EPI_ISL_1838227, EPI_ISL_1838229, EPI_ISL_1838230, EPI_ISL_1838234, EPI_ISL_1838235, EPI_ISL_1838237, EPI_ISL_1838241, EPI_ISL_1838253, EPI_ISL_1838256, EPI_ISL_1838259, EPI_ISL_1838260, EPI_ISL_1838262, EPI_ISL_1838264, EPI_ISL_1838265, EPI_ISL_1838267, EPI_ISL_1838271, EPI_ISL_1838274, EPI_ISL_1838277, EPI_ISL_1838280, EPI_ISL_1838282, EPI_ISL_1838285, EPI_ISL_1838292, EPI_ISL_1838295, EPI_ISL_1838309, EPI_ISL_1838312, EPI_ISL_1838315, EPI_ISL_1838318, EPI_ISL_1838322, EPI_ISL_1838325, EPI_ISL_1838327, EPI_ISL_1838333, EPI_ISL_1838337, EPI_ISL_1838338, EPI_ISL_1838345 | | | |
| see above | The National Centre for Cell Science | CSIR-Centre for Cellular and Molecular Biology-INSACOG | Dhiraj Paul, Mohak P Gujare, Shivang P. Bhanushali, Mitali Inamdar, Sonal Manik Chavan, Manoj Kumar Bhat, Ajay Pillai, INSACOG Consortium team, Yogesh Shouche,Payel Mukherjee,Lamuk Zaveri,Tulasi Nagabandi, Ara Sreenivas,Valli Nagalakshmi Undamatla,Shreekant Verma, Amareshwar Vodapalli ,Blessy B John,Viswagithe S L,B Himasri,Onkar Kulkarni,Sofia Banu,Archana Bharadwaj Siva,Sharath Chandra Thota,Karthik Bharadwaj Tallapaka,Rakesh K Mishra,Divya Tej Sowpati |
| EPI_ISL_1838348, EPI_ISL_1838350, EPI_ISL_1838356, EPI_ISL_1838359, EPI_ISL_1838364, EPI_ISL_1838366, EPI_ISL_1838367, EPI_ISL_1838369, EPI_ISL_1838377, EPI_ISL_1838380, EPI_ISL_1838385, EPI_ISL_1838392, EPI_ISL_1838393, EPI_ISL_1838400, EPI_ISL_1838410, EPI_ISL_1838413, EPI_ISL_1838414, | | | |

| | | | |
|---|---|---|---|
| EPI_ISL_1838418, EPI_ISL_1838430, EPI_ISL_1838446, EPI_ISL_1838448, EPI_ISL_1838452, EPI_ISL_1838454, EPI_ISL_1838455, EPI_ISL_1838456, EPI_ISL_1838457, EPI_ISL_1838459, EPI_ISL_1838460, EPI_ISL_1838464, EPI_ISL_1838466, EPI_ISL_1838467, EPI_ISL_1838468, EPI_ISL_1838471, EPI_ISL_1838472, EPI_ISL_1838473, EPI_ISL_1838477, EPI_ISL_1838489, EPI_ISL_1838494, EPI_ISL_1838502, EPI_ISL_1838520, EPI_ISL_1838522, EPI_ISL_1838526, EPI_ISL_1838543, EPI_ISL_1838545, EPI_ISL_1838546, EPI_ISL_1838547, EPI_ISL_1838548, EPI_ISL_1838550, EPI_ISL_1838551, EPI_ISL_1838552, EPI_ISL_1838553, EPI_ISL_1838577, EPI_ISL_1838578, EPI_ISL_1838580, EPI_ISL_1838581, EPI_ISL_1838583, EPI_ISL_1838588, EPI_ISL_1838593, EPI_ISL_1838594, EPI_ISL_1838604, EPI_ISL_1838605, EPI_ISL_1838610, EPI_ISL_1838613, EPI_ISL_1838615, EPI_ISL_1838616, EPI_ISL_1838617, EPI_ISL_1838618, EPI_ISL_1838619, EPI_ISL_1838620, EPI_ISL_1838621, EPI_ISL_1838622, EPI_ISL_1838623, EPI_ISL_1838626, EPI_ISL_1838629, EPI_ISL_1838631, EPI_ISL_1838635, EPI_ISL_1838636, EPI_ISL_1838638, EPI_ISL_1838640, EPI_ISL_1838650, EPI_ISL_1838652, EPI_ISL_1838654, EPI_ISL_1838655, EPI_ISL_1838658, EPI_ISL_1838663, EPI_ISL_1838664, EPI_ISL_1838668, EPI_ISL_1838669, EPI_ISL_1838670, EPI_ISL_1838675, EPI_ISL_1838694, EPI_ISL_1838696, EPI_ISL_1838697, EPI_ISL_1838699, EPI_ISL_1838708, EPI_ISL_1838711, EPI_ISL_1838712, EPI_ISL_1838716, EPI_ISL_1838718, EPI_ISL_1838719, EPI_ISL_1838722, EPI_ISL_1838725, EPI_ISL_1838729, EPI_ISL_1838730, EPI_ISL_1838731, EPI_ISL_1838732, EPI_ISL_1838733, EPI_ISL_1838736, EPI_ISL_1838737, EPI_ISL_1838738, EPI_ISL_1838739, EPI_ISL_1838740, EPI_ISL_1838741, EPI_ISL_1838744, EPI_ISL_1838745, EPI_ISL_1838746, EPI_ISL_1838748, EPI_ISL_1838750, EPI_ISL_1838755, EPI_ISL_1838761, EPI_ISL_1838762, EPI_ISL_1838763, EPI_ISL_1838764, EPI_ISL_1838766, EPI_ISL_1838768, EPI_ISL_1838769, EPI_ISL_1838770, EPI_ISL_1838771, EPI_ISL_1838772, EPI_ISL_1838774, EPI_ISL_1838775, EPI_ISL_1838776, EPI_ISL_1838777, EPI_ISL_1838778, EPI_ISL_1838779, EPI_ISL_1838780, EPI_ISL_1838785, EPI_ISL_1838786, EPI_ISL_1838787, EPI_ISL_1838788, EPI_ISL_1838789, EPI_ISL_1838790, EPI_ISL_1838795, EPI_ISL_1838798, EPI_ISL_1838799, EPI_ISL_1838802, EPI_ISL_1838803, EPI_ISL_1838804, EPI_ISL_1838806, EPI_ISL_1838809, EPI_ISL_1838810, EPI_ISL_1838811, EPI_ISL_1838812, EPI_ISL_1838813, EPI_ISL_1838814, EPI_ISL_1838815, EPI_ISL_1838817, EPI_ISL_1838818, EPI_ISL_1838819, EPI_ISL_1838820, EPI_ISL_1838821, EPI_ISL_1838822, EPI_ISL_1838823, EPI_ISL_1838824, EPI_ISL_1838825, EPI_ISL_1838827, EPI_ISL_1838828, EPI_ISL_1838831, EPI_ISL_1838832, EPI_ISL_1838833, EPI_ISL_1838834, EPI_ISL_1838835, EPI_ISL_1838836, EPI_ISL_1838837, EPI_ISL_1838838, EPI_ISL_1838839, EPI_ISL_1838841, EPI_ISL_1838842, EPI_ISL_1838843, EPI_ISL_1838844, EPI_ISL_1838845, EPI_ISL_1838846, EPI_ISL_1838848, EPI_ISL_1838849, EPI_ISL_1838851, EPI_ISL_1838852, EPI_ISL_1838853, EPI_ISL_1838854, EPI_ISL_1838855, EPI_ISL_1838856, EPI_ISL_1838857, EPI_ISL_1838858, EPI_ISL_1838859, EPI_ISL_1838860, EPI_ISL_1838861, EPI_ISL_1838879, EPI_ISL_1838882, EPI_ISL_1838900, EPI_ISL_1838914 | | | |
| see above | CSIR-Centre for Cellular and Molecular Biology | CSIR-Centre for Cellular and Molecular Biology-INSACOG | Payel Mukherjee,Lamuk Zaveri,Tulasi Nagabandi, Ara Sreenivas,Shreekant Verma, Amareshwar Vodapalli ,Blessy B John,Viswagithe S L,B Himasri,Valli Nagalakshmi Undamatla,Onkar Kulkarni,Sofia Banu,Archana Bharadwaj Siva,Sharath Chandra Thota,Karthik Bharadwaj Tallapaka,Rakesh K Mishra,Divya Tej Sowpati |
| EPI_ISL_1841236, EPI_ISL_1841237, EPI_ISL_1841238, EPI_ISL_1841239, EPI_ISL_1841240, EPI_ISL_1841241, EPI_ISL_1841242, EPI_ISL_1841243, EPI_ISL_1841244, EPI_ISL_1841245, EPI_ISL_1841246, EPI_ISL_1841248, EPI_ISL_1841249, EPI_ISL_1841250, EPI_ISL_1841251, EPI_ISL_1841252, EPI_ISL_1841253, EPI_ISL_1841254, EPI_ISL_1841255, EPI_ISL_1841256, EPI_ISL_1841257, EPI_ISL_1841258, EPI_ISL_1841259, EPI_ISL_1841260, EPI_ISL_1841261, EPI_ISL_1841262, EPI_ISL_1841263, EPI_ISL_1841265, EPI_ISL_1841266, EPI_ISL_1841267, EPI_ISL_1841268, EPI_ISL_1841269, EPI_ISL_1841270 | | | |
| see above | ICMR-National Institute of Virology - INSACOG | NIV Influenza | Dr. Varsha Potdar |
| EPI_ISL_953400, EPI_ISL_981014, EPI_ISL_981015, EPI_ISL_981027, EPI_ISL_995306 | National Public Health Laboratory, National Centre for Infectious Diseases | National Public Health Laboratory, National Centre for Infectious Diseases | Tze Minn Mak, Zhenyang Zhou, Lin Cui, Raymond Tzer Pin Lin |
